# Immunological profiles in Lynch syndrome colorectal cancers are not specific to mismatch repair gene defects

**DOI:** 10.1101/2024.08.27.24311855

**Authors:** Noah C. Helderman, Marieke E. IJsselsteijn, Madalina Cabuta, Manon van der Ploeg, Tom van Wezel, Aysel Ahadova, Matthias Kloor, Hans Morreau, Maartje Nielsen, Noel F.C.C. de Miranda

## Abstract

**Background and aims:** Colorectal carcinomas (CRCs) in patients with Lynch syndrome (LS) exhibit heightened immunogenicity due to mismatch repair deficiency (MMR-d), often resulting in favorable responses to T cell immune checkpoint therapies. Recent studies indicate that the phenotype and genotype of LS-associated CRCs vary depending on the specific MMR gene mutated. Here, we investigated whether the immune profiles of LS-associated CRCs differ based on the MMR gene defects.

**Methods:** Tissue material from 18 *MLH1-,* 16 *MSH2-,* 40 *MSH6-, and* 23 *PMS2-*mutated CRCs and 35 sporadic MMR-d CRCs were included in the study. Imaging mass cytometry (IMC) analysis, along with targeted multiplex immunofluorescence imaging (mIF) and immunohistochemistry, were applied to examine the tumor immune microenvironment, including Human Leukocyte Antigen (HLA) class I and programmed death-ligand 1 (PD-L1) expression.

**Results:** Unsupervised hierarchical clustering of cell phenotypes identified by IMC, followed by mIF validation, revealed comparable lymphoid and myeloid cell infiltration levels across CRCs from all MMR groups. Infiltrating T cell levels negatively correlated with the number of mutations at coding microsatellite sequences, particularly in *MLH1-*mutated CRCs. HLA class I defects were observed in 76% of all CRCs. These defects were more frequently accompanied by β2M defects in hereditary MMR-d CRCs (67%) compared to sporadic MMR-d CRCs (37%), and did not associate with the number of γδ T cells, which were present in CRCs from all MMR groups. PD-L1 expression in tumor cells was only detected in 8% of all CRCs.

**Conclusion:** Our findings illustrate that, from an immunological perspective, there is no evidence of differing immunogenic features across MMR defects. This is important to consider when developing preventive vaccine strategies and evaluating immunotherapy for LS patients and those with MMR-d CRCs.

## Introduction

Lynch syndrome (LS) is recognized as one of the most prevalent inherited cancer syndromes, primarily conferring a predisposition to colorectal cancer (CRC) and endometrial cancer. It arises in carriers of pathogenic variants in one of the mismatch repair (MMR) genes, including *MLH1* (OMIM #609310), *MSH2* (OMIM #120435), *MSH6* (OMIM #614350), and *PMS2* (OMIM #614337), or more rarely from an *EPCAM* (OMIM #613244) deletion that lies upstream of *MSH2*.^1^

In general, CRCs from LS patients are highly immunogenic due to the widespread accumulation of mutations, particularly nucleotide insertions and deletions at microsatellite sequences, due to MMR deficiency (MMR-d).^1^ Insertions and deletions in the coding genome are thought to be particularly immunogenic due to the generation of frameshift proteins that are new to the host’s immune system and, therefore, evoke anti-tumor immune reactivity.^2^

The remarkable immunogenicity of LS-associated CRCs is evident through pronounced infiltration of (cytotoxic) T cells, the frequent observation of immune evasion events, and immune cell reactivity to frameshift proteins in LS-associated CRCs.^3–14^ Capitalizing on this, patients with LS-associated CRCs or sporadic MMR-d CRCs generally respond better to immune checkpoint blockade than those with MMR-proficient CRCs.^15–19^ Furthermore, this population may benefit from preventive cancer vaccine development, as certain frameshift proteins are shared across most MMR-d CRCs due to the positive selection of underlying driver mutations. ^20, 21^

Recent studies demonstrated that LS presents as a highly heterogeneous disease, encompassing variation in both phenotypic aspects such as CRC risks^22–30^, and genotypic factors, including the type of somatic mutations (e.g., single-nucleotide variants, insertions, deletions) and the genes affected by these mutations.^31–37^ These disparities are likely influenced by the underlying MMR defect, among other contributing factors, and have prompted whether LS should be redefined as a collection of distinct gene-related syndromes.^38, 39^

We recently observed a reduced frequency of MMR-d signature-associated insertion and deletion mutations in *MSH6*-mutated CRCs compared to other LS-associated CRCs.^36, 37^ Given that these mutations typically underlie the observed immune responses in CRCs, our goal was to conduct a comprehensive characterization of the immune profiles in LS-associated CRCs to determine whether those are influenced by specific MMR gene defects.

## Methods

### Ethical statement

The Medical Ethical Committee of Leiden, The Hague, Delft (protocol P17.098) approved this study. Patient samples were managed in accordance with medical ethical guidelines outlined in the Code of Conduct for the responsible use of human tissue in the context of health research, as established by the Federation of Dutch Medical Scientific Societies. Informed consent was obtained from patients to utilize both tissue and data. Patients or the public were not involved in the design, or conduct, or reporting, or dissemination plans of our research

### Patients and samples

Coded or anonymized formalin-fixed, paraffin-embedded (FFPE) tumor tissue blocks were obtained from the Department of Pathology of the Leiden University Medical Centre (Leiden, The Netherlands) and via the Dutch Pathology Registry (PALGA; reference LZV2022-68), respectively. The tumor tissue that was included originated from CRCs of 18 *MLH1*, 16 *MSH2*, 40 *MSH6*, and 23 *PMS2* variant carriers. Moreover, tumor tissue blocks from 35 patients with sporadic MMR-d CRC due to *MLH1* promotor hypermethylation (*MLH1-*PM) were included. Clinical and molecular characteristics of patients and samples were obtained from pathology reports and patient records.

### Immunohistochemistry

See **Supplementary Table 1** for providers and relevant (user) details of all materials used in this study, including antibody clones.

#### Immunodetection

FFPE tumor tissue blocks were cut into 4μm sections and placed on silane-coated glass slides (VWR, Radnor, PA, USA). Immunohistochemical (IHC) detection of human leukocyte antigen (HLA) class I [HCA2 and HC10 clones], β2-microglobulin (β2M) [EPR21752-214 clone], programmed death-ligand 1 (PD-L1) [E1L3N clone], and TCR δ [H-41], was performed as described previously.^10^ In short, tissue sections were deparaffinized and rehydrated using xylene and decreasing ethanol concentrations, respectively. Endogenous peroxidase was blocked using 0.3% hydrogen/peroxidase methanol solution (Merck Millipore, Burlington, MA, USA) and heat-mediated antigen retrieval was achieved by boiling the sections in either sodium-citrate (0.1M, pH 6) or Tris-EDTA (10/1mM, pH 9) buffer, following which the sections were cooled and incubated overnight with a primary antibody (**Supplementary Table 1**). Primary antibody binding was detected through incubation with BrightVision poly-horseradish peroxidase solution (Immunologic, Duiven, The Netherlands) and DAB+ chromogen (DAKO, Agilent Technologies, Santa Clara, CA, USA). Sections were counterstained with hematoxylin (Thermo Fisher Scientific, Waltham, MA, USA).

#### Scoring and quantitative analysis

An internal positive control (stromal cell staining) was used to score HLA class I, β2M, and PD-L1 expression in tumor cells (**Supplementary Figure 1**). Negative controls comprised 4μm tonsil tissue incubated with 1% bovine serum albumin (BSA)/PBS solution instead of primary antibodies during the procedure.

Quantitative analysis of γδ T cells in whole slide scans of tumor tissue was carried out using QuPath v0.4.3.^40^ Initially, cell segmentation was performed utilizing Hematoxylin staining to identify cell nuclei. Subsequently, the positivity of the TCR V γ 3 marker in the cells was determined based on DAB intensity. Normalization was conducted to account for tissue area variations, and the resulting cell counts were reported as cells/mm².

### Imaging mass cytometry

To identify immune cell phenotypes of interest, we applied a 40-marker IMC panel that has previously been designed and optimized^41^, on eight tumors per MMR group. This approach allowed the analysis of both immune cell frequencies and the spatial context of immune cells. Antibody-metal conjugation, IMC immunodetection/acquisition, and downstream data analyses were performed as described previously by our group and will be summarized.^41–43^

#### Metal conjugation of antibodies

Conjugation of carrier-free IgG antibodies to purified Ianthanide metals was performed using the Maxpar antibody labeling kit (Fluidigm, San Francisco, CA, USA), following which antibodies were eluted in 50μL antibody stabilizer solution (Candor Bioscience, Wangen im Allgäu, Germany) with 0,05% sodium azide and 50μl W-buffer (Fluidigm). Antibody performance after conjugation was assessed by IHC on 4μm tonsil tissue. The marker panel included lineage, functional/activation, and structural markers (**Supplementary Table 1**).

#### Immunodetection

A tissue microarray containing two tissue cores (diameter 1.5mm) per CRC was constructed from FFPE tumor tissue blocks using the TMA Master (3DHISTECH Kft, Budapest, Hungary). Tonsil, sigmoid colon, and placental tissue were included as references. Regions of interest were selected on hematoxylin and eosin stains. A 4μm section was cut from the tissue microarray, deparaffinized, and rehydrated using xylene and decreasing ethanol concentrations. Heat-mediated antigen retrieval was achieved by boiling the section in sodium citrate (0.1M, pH 6), after which the section was incubated with Superblock solution (Thermo Fisher Scientific) to minimize non-specific antibody binding. Following washes in PBS (supplemented with 0.05% tween and 1% BSA), the section was incubated with antibody-metal conjugates using the conditions described in **Supplementary Table 1**.

#### Data acquisition

The Hyperion mass cytometry imaging system (Fluidigm) was autotuned using a 3-element tuning slide (Fluidigm), with a minimal detection of 1500 mean duals of 175Lu being required as an extra threshold for successful tuning. For each tissue core, an area of 1000x1000μm was ablated at 200Hz. Raw data was exported as MCD files and visualized with MCD viewer (Fluidigm).

#### Image enhancement, semi-automated background removal, and creation of single cell masks

MCD files were converted into tiff files using MCD viewer, following which images were enhanced (removal of outliers with values <1^st^ or >99^th^ percentile) using MATLAB (MathWorks, Natick, MA, USA). Ilastik’s random forest classifier^44^ was trained to distinguish background from real signal for every marker (exporting data as binary expression maps with background pixels set to 0 and real pixels set to 1) and to create probability masks for nuclei (based on DNA signal), tumor membranes (based on keratin signal) and stromal membranes (based on vimentin signal).^42^ Using these probability masks, single-cell masks were created for all samples with CellProfiler^45^, which were validated through visual comparison with the original IMC images.

#### Single-cell clustering and phenotype calling

Single-cell masks of each sample and binary expression maps of each marker were loaded into ImaCytE^46^, in which the relative frequencies of positive pixels per cell were visualized and exported as single-cell FCS files. The latter were analyzed by t-distributed stochastic neighborhood embedding (t-SNE) in Cytosplore^47^, in which Mean-shift clustering was used to group visual neighborhoods in the t-SNE embedding. The resulting groups were assigned a phenotype name based on the expression of lineage-specific markers (**Supplementary Table 2**) and were loaded back into ImaCytE for visual validation.

### Multiplex immunofluorescence imaging

To further evaluate the immune cell phenotypes of interest as identified by IMC, we next applied mIF imaging using selected panels of antibodies on the entire set of tumors (**Supplementary Table 1**). For the assessment of T cells, simultaneous detection of CD4, CD8, CD103, granzyme B (GZMB), and PD-1 was applied, whereas a triple mIF panel targeting CD204, HLA-DR, and CD15 was used to evaluate myeloid cell infiltration.

#### Immunodetection

The process involved deparaffinization, endogenous peroxidase blocking, and heat-induced epitope retrieval steps akin to those described in the IHC procedure, using 4μm FFPE sections. Next, an Opal amplified detection process was applied. To minimize non-specific antibody binding, slides were incubated with Superblock solution. Primary antibody incubation lasted one hour, after which BrightVision Goat Anti-Mouse/Rabbit IgG HRP (Immunologic) was applied as the secondary antibody. After each antibody application, different Opal fluorophores (690, 650 620, 570, 520; Akoya Biosciences, Marlborough, MA, USA) were used to visualize multiple markers within the same tissue section. Microwave treatment was then conducted to remove primary and secondary antibodies and other non-specific staining and reduce tissue autofluorescence. Following each incubation period, slides were washed three times with a 0.05% Tween in PBS solution. An additional step involving DAPI incubation was carried out to stain the nuclei. Tissue slides were mounted using ProLong Gold Antifade Reagent (Cell Signaling Technologies, MA, USA). Tonsil slides were included as negative controls. These were subjected to incubation with 1% bovine serum albumin (BSA)/PBS in place of the primary antibody.

#### Data acquisition

mIF images were captured using the Vectra 3.0 Automated Quantitative Pathology Imaging system (Akoya Biosciences). Whole slide scans for DAPI stains were performed at 4x magnification, and approximately four areas of interest were designated per sample using Phenochart Software (Akoya Biosciences). The multispectral images (MSIs) were obtained at 20x magnification, and spectral separation of dyes was carried out using the Inform Cell Analysis Software (Akoya Biosciences). The spectral library was created using single-stain mIF slides.

#### Phenotype calling and cell counting

The PENGUIN tool was employed for image normalization, rescaling all images and markers between 0 and 1.^48^ Subsequently, a two-step denoising process was applied, wherein a minimal signal threshold was set for each marker, followed by percentile normalization. Cell segmentation masks were created from the normalized images using CellProfiler.^45^ Initially, nuclei were defined using the DNA images, and membranes were added using the membrane markers from the dataset. For each cell outlined by CellProfiler, the mean intensity of each marker was calculated. Employing the FlowSOM R-package (version 2.6.0)^49^, all cells were clustered based on mean marker intensity (FlowSOM settings: xdim = 5, ydim = 4, nClus = 5). Cluster evaluation involved visual inspection, with merging occurring for clusters exhibiting similar marker profiles and guided by prior knowledge of marker biology. The final clusters were remapped onto the images and validated by comparison with raw data.

#### Correlation with coding microsatellite mutations

To evaluate the potential influence of the molecular background on the infiltrating T cell levels, we correlated the T cell counts from the current study with the number of cMS mutations per tumor (max. 20 cMS tested per tumor). The latter data is considered to be a measure of the degree of MSI^50^ and was obtained from Helderman et al.^37^, where we focused on the same cohort of tumors.

### Statistical analysis

Statistical analyses were conducted using RStudio (Team R, Integrated Development for R, Boston, MA, 2020). Continuous variables are expressed as mean (standard deviation, SD) or median (interquartile range, IQR) and were compared using either the (un)paired t-test or Mann-Whitney U test for two groups and the ordinary one-way ANOVA or Kruskal-Wallis test for more than two groups. Categorical variables are presented as proportions and were compared using Pearson’s χ² test or Fisher’s exact test for two categories, and Pearson’s χ² test for more than two categories. Correlations were assessed using Pearson correlation. When applicable, raw *P* values were adjusted for the number of comparisons and outcomes using the Benjamini & Hochberg correction for multiple testing. All *P* values reported in this article are two-tailed and considered statistically significant at *P*<0.05. Cell counts were visualized in box and whisker plots displaying all data points (one point per tumor sample) and in heatmaps where phenotypes and MMR groups underwent unsupervised hierarchical clustering, using RStudio (Team R, Integrated Development for R, Boston, MA, 2020) and the ComplexHeatmap package.^51^

## Results

An overview of the clinical and histological characteristics of each MMR group, encompassing 18 *MLH1*-, 16 *MSH2*-, 40 *MSH6*-, and 23 *PMS2*-mutated CRCs, along with 35 sporadic MMR-d CRCs due to *MLH1*-PM, is provided in **Supplementary Table 3**. A full description of all available clinical and histological characteristics for each analyzed CRC (including tumor IDs) is presented in **Supplementary Table 4**. Of note, *MSH6*-mutated CRCs were diagnosed at higher ages and more often located distally than other LS-associated CRCs.

### Immune profiles are not specific to MMR mutations

To examine the immune landscape of LS-associated CRCs, we initially applied a 40-marker IMC panel to eight tumor tissues per MMR group (with successful data acquisition for all but one *PMS2*-mutated CRC, tumor ID PMS2_08).^41^ Based on this panel; we were able to identify five cancer, eleven lymphoid (including eight T cell phenotypes), seven myeloid, and four stromal cell phenotypes (**Supplementary Table 2**). The median cell count per MMR group for each identified phenotype through IMC is detailed in **Supplementary Table 5**.

Unsupervised hierarchical clustering of the IMC phenotypes did not identify clusters uniquely associated with selected MMR group(s), except for *MSH6*-mutated CRCs, which were generally characterized by a lower relative frequency of T cells (**Figure 1A**). In line with the latter, minor distinctions were observed in the levels of (CD4^+^; CD8^+^) T cells, which tended to be higher in *MLH1*- mutated CRCs, *MLH1*-PM CRCs, and a subset of *PMS2*-mutated CRCs compared to *MSH2*-mutated CRCs and *MSH6*-mutated CRCs (**Supplementary Figure 2**, **Supplementary Table 5**). A similar trend was noted for CD8^+^ T cells expressing GZMB (a serine protease secreted by activated CD8^+^ T cells^52^), PD-1 (an inhibitory checkpoint molecule expressed by activated/primed T cells^53^), and CD103 (a mediator of adhesion and tissue retention of CD8^+^ T cells^54, 55^) (**Supplementary Figure 3**, **Supplementary Table 5**). In the myeloid cell compartment, granulocytes (CD15^+^ cells) also appeared more prevalent in *MLH1*-mutated, *MLH1*-PM, and *PMS2*-mutated CRCs compared to *MSH2*- and *MSH6*-mutated CRCs (**Supplementary Figure 2**, **Supplementary Table 5**). Conversely, the count of CD204^+^ macrophages was slightly higher in *MSH2*-mutated CRCs (**Supplementary Figure 2**, **Supplementary Table 5**).

**Figure 1.**
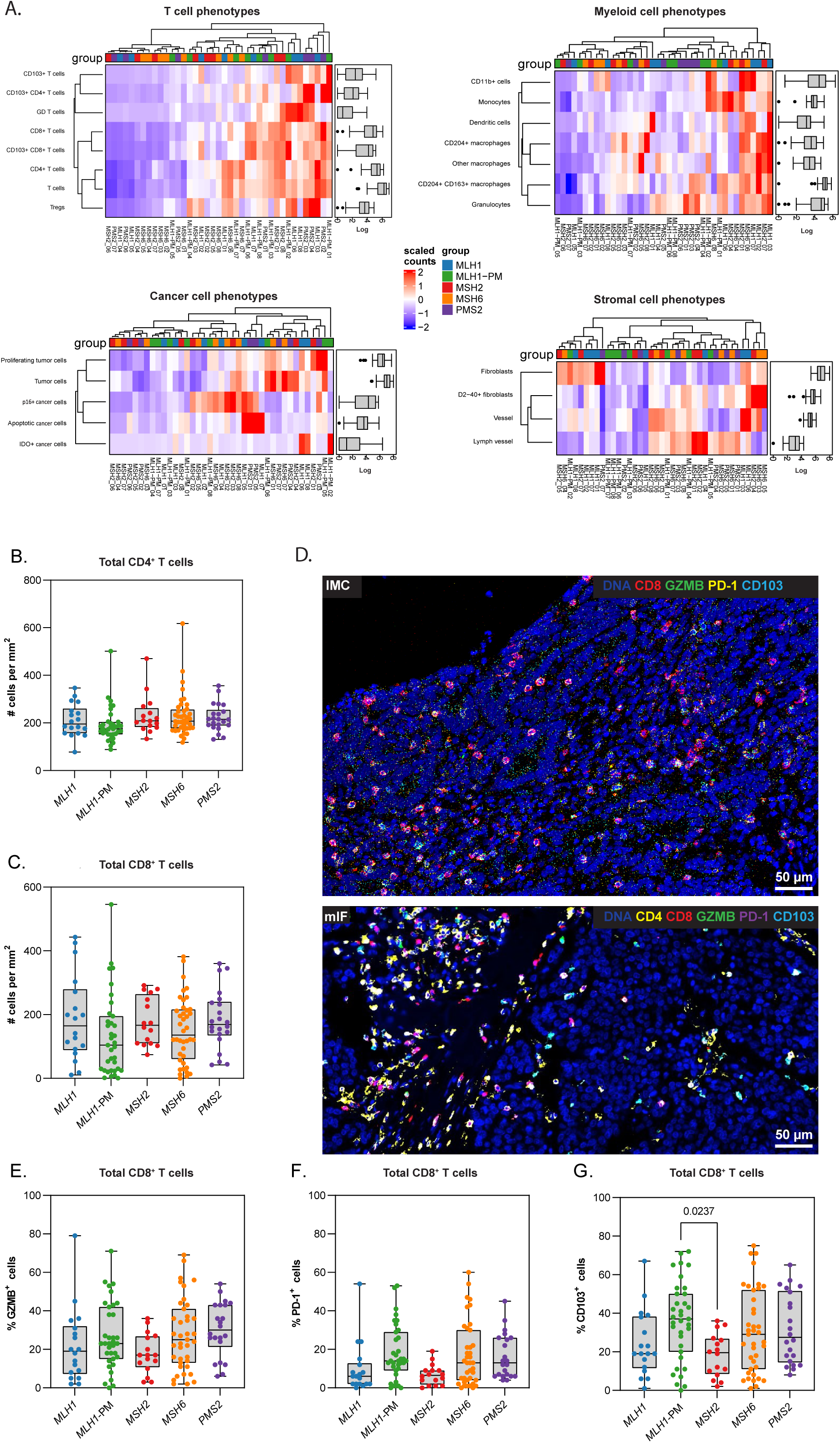
Immune profiles are not specified by the MMR gene defect. [**A**] Unsupervised hierarchical clustering of the phenotypes identified through IMC revealed considerable heterogeneity amongst the CRCs, but did not identify clusters specifically associated with selected MMR group(s). The heatmap showcasing lymphoid cell phenotypes only features T cell phenotypes, excluding B cells, plasma cells/plasmablasts, and innate lymphoid cells. The tumor IDs are shown on the x-axis. The overall number of [**B**] CD4^+^ T cells (all CD4^+^ T cell phenotypes combined) and [**C**] CD8^+^ T cells (all CD8^+^ T cell phenotypes combined) did not differ between the MMR groups. [**D**] Illustrative IMC image showing detection of DNA (dark blue), CD8 (red), GZMB (green), PD-1 (yellow), and CD103 (light blue), along with an illustrative mIF image showing detection of DNA (dark blue), CD4 (yellow), CD8 (red), GZMB (green), PD-1 (purple), and CD103 (light blue). The proportion of CD8^+^ T cells expressing [**C**] GZMB, [**D**] PD-1, and [**E**] CD103 did not vary between the MMR groups. Cell counts are visualized in box and whisker plots displaying all data points (one point per tumor sample). *CRC, colorectal cancer; GZMB, granzyme B; IMC, imaging mass cytometry; mIF, multiplex immunofluorescence; MLH1-PM, MLH1 promotor hypermethylation; MMR, mismatch repair; PD-1, programmed death 1*.

Based on the previous observations, we next applied targeted mIF and IHC with selected antibody panels on full slides of the entire set of tumors to validate the IMC results of the phenotypes discussed in the previous paragraph. The median cell count per MMR group for the phenotypes observed through mIF and IHC is presented in **Supplementary Table 6**.

Firstly, we evaluated CD4^+^ and CD8^+^ T cells, including their CD103/GZMB/PD-1 status, using mIF. Based on the mIF dataset, the median density of CD4^+^ T cells (**Figure 1B**) and CD8^+^ T cells (**Figure 1C**) was consistent across CRCs in each MMR group (**Figure 1D**). Similarly, the proportion of CD4^+^ T cells and CD8^+^ T cells expressing GZMB (**Figure 1E**), PD-1 (**Figure 1F**), and/or CD103 (**Figure 1G**) showed only modest variation between CRCs from each MMR group, thereby suggesting that T cell infiltration and activation does not seem to be influenced by a specific MMR defect.

Secondly, we analyzed CD204^+^ macrophages and granulocytes through mIF. Similar to the lack of evident differences regarding the lymphoid phenotypes, we noted comparable levels of CD204^+^ macrophage and granulocyte infiltration across CRCs in every MMR group (**Supplementary Figure 4**).

To determine if the comparable immune profiles of the different MMR groups observed through mIF were also mirrored by similar immune evasive events in various LS-associated CRCs, we next examined HLA class I and PD-L1 expression by IHC across the entire set of tumors. HLA class I defects were observed in 73% of all MMR-d CRCs (**Figure 2A**), consistent with previous studies.^9, 10^ Total loss of HLA class I expression (absent membranous detection of both HCA2 and HC10 clones) was seen in 53% of cases, while partial loss (absent membranous detection of either HCA2 or HC10) was noted in 20% of cases. Among the CRCs with any HLA class I defect(s) (total or partial loss), aberrant β2M expression was seen less frequently (*P*=<0.001) in sporadic *MLH1-*PM CRCs (37%) versus hereditary *MLH1*- (85%)*, MSH2-* (58%)*, MSH6-* (70%) and *PMS2-*mutated (53%) CRCs (**Figure 2B**). PD-L1 expression could be detected in tumor cells in a minority of *MLH1-*PM CRCs (20%), *MSH2-* (25%), *MSH6-* (17%), and *PMS2-*mutated (4%) CRCs, accounting for 8% of all CRCs (**Figure 2C**).

**Figure 2.**
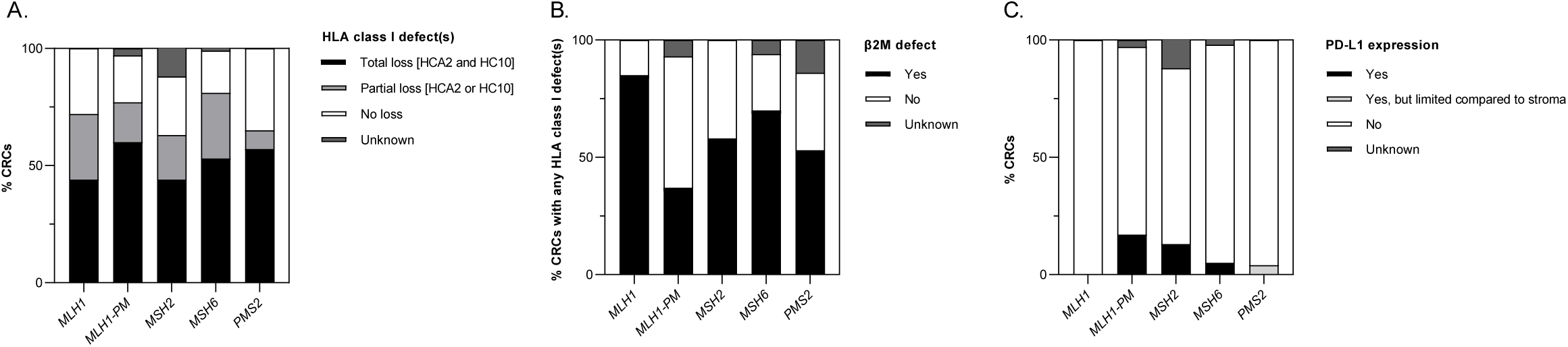
Frequent HLA class I defects in CRCs from all MMR groups. [**A**] Percentages of CRCs showing total loss (absent membranous detection of both HCA2 and HC10 clones) or partial loss (absent membranous detection of either HCA2 or HC10) of HLA class I expression per MMR group. [**B**] Percentage of CRCs with any HLA class I defect(s) (total or partial loss) showing aberrant β2M expression per MMR group. [**C**] Percentage of CRCs in which PD-L1 expression was detected, stratified by MMR group. Evaluation was performed by IHC. Stromal cell staining was used as internal positive control. *β2M, β2-microglobulin; HLA, human leukocyte antigen; IHC, immunohistochemistry; MLH1- PM, MLH1 promotor hypermethylation; MMR, mismatch repair; PD-L1, programmed death-ligand 1*.

We then assessed the presence of γδ T cells (GD T cells; CD3^+^CD45^+^GD^+^ cells) in the entire set of tumors through conventional IHC for the TCR δ [H-41] clone since γδ T cells were recently discovered to be involved in responses to immune checkpoint blockade in MMR-d CRCs with HLA class I defects^56^. As per the IMC data (**Supplementary Table 5**), γδ T cells were identified in varying degrees in CRCs from all MMR groups. IHC analysis of the entire cohort validated γδ T cells to be present in the majority (84%) of CRCs, with the median γδ T cell count being comparable in each MMR group (**Figure 3A-B**). The γδ T cell count did not associate with the tumors’ HLA class I status (**Figure 3C**).

**Figure 3.**
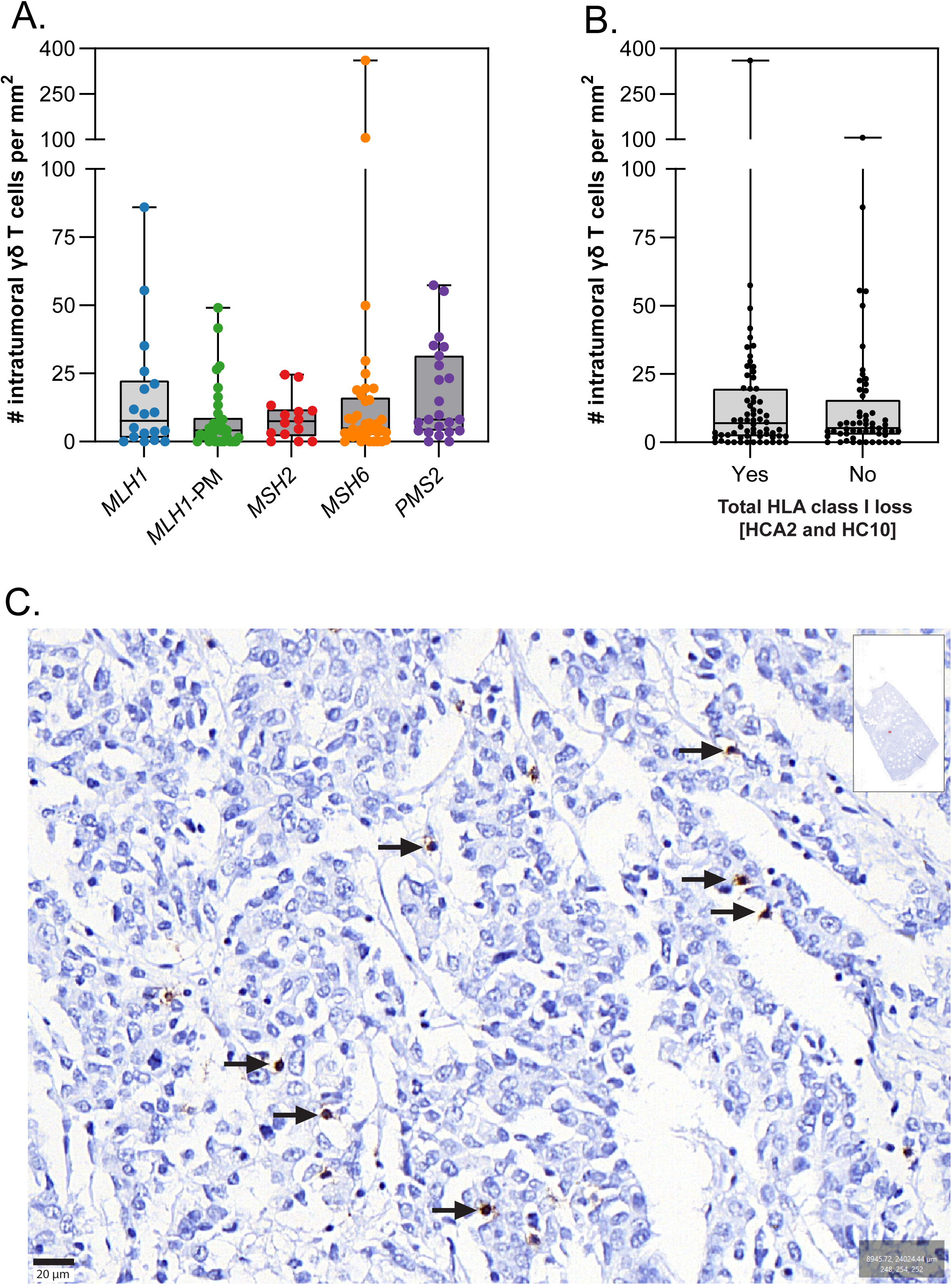
γδ T cells are present in the majority of LS-associated CRCs independent of the MMR and HLA class I status. [**A**] The median number of γδ T cells was comparable between CRCs from every MMR group and [**B**] was independent of the HLA class I status (total loss [HCA2 and HC10]). [**C**] Illustrative IHC image showing γδ T cells (indicated by arrows) in a *MSH6-*mutated CRC. *HLA, human leukocyte antigen; LS, Lynch syndrome; MMR, mismatch repair*.

### CD8^+^ T cell levels negatively correlate with cMS mutations in *MLH1-*mutated CRCs

Finally, we correlated the T cell counts from the current study with the number of cMS mutations obtained from Helderman et al.^37^ to evaluate the potential influence of the molecular background on the infiltrating T cell levels. Negative correlations were observed between the number of cMS mutations and the number of CD4^+^ T cells (all CD4^+^ T cell phenotypes combined) when considering all CRCs (Pearson’s r −0.221; *P*=0.013) or when specifically considering *MLH1-*PM CRCs (Pearson’s r −0.370; *P*=0.029) (**Figure 4A-B**, **Supplementary Figure 5**). Likewise, a negative correlation was identified between the number of cMS mutations and the number of CD8^+^ T cells (all CD8^+^ T cell phenotypes combined) when considering the *MLH1*-mutated CRCs only (Pearson’s r −0.540; *P*=0.021) (**Figure 4C-D**, **Supplementary Figure 5**). The HLA class I status (total loss) was unaffected (*P*=0.125) by whether tumors had fewer than the median number (=13) of cMS mutations or equal to or more than the median number of cMS mutations.

**Figure 4.**
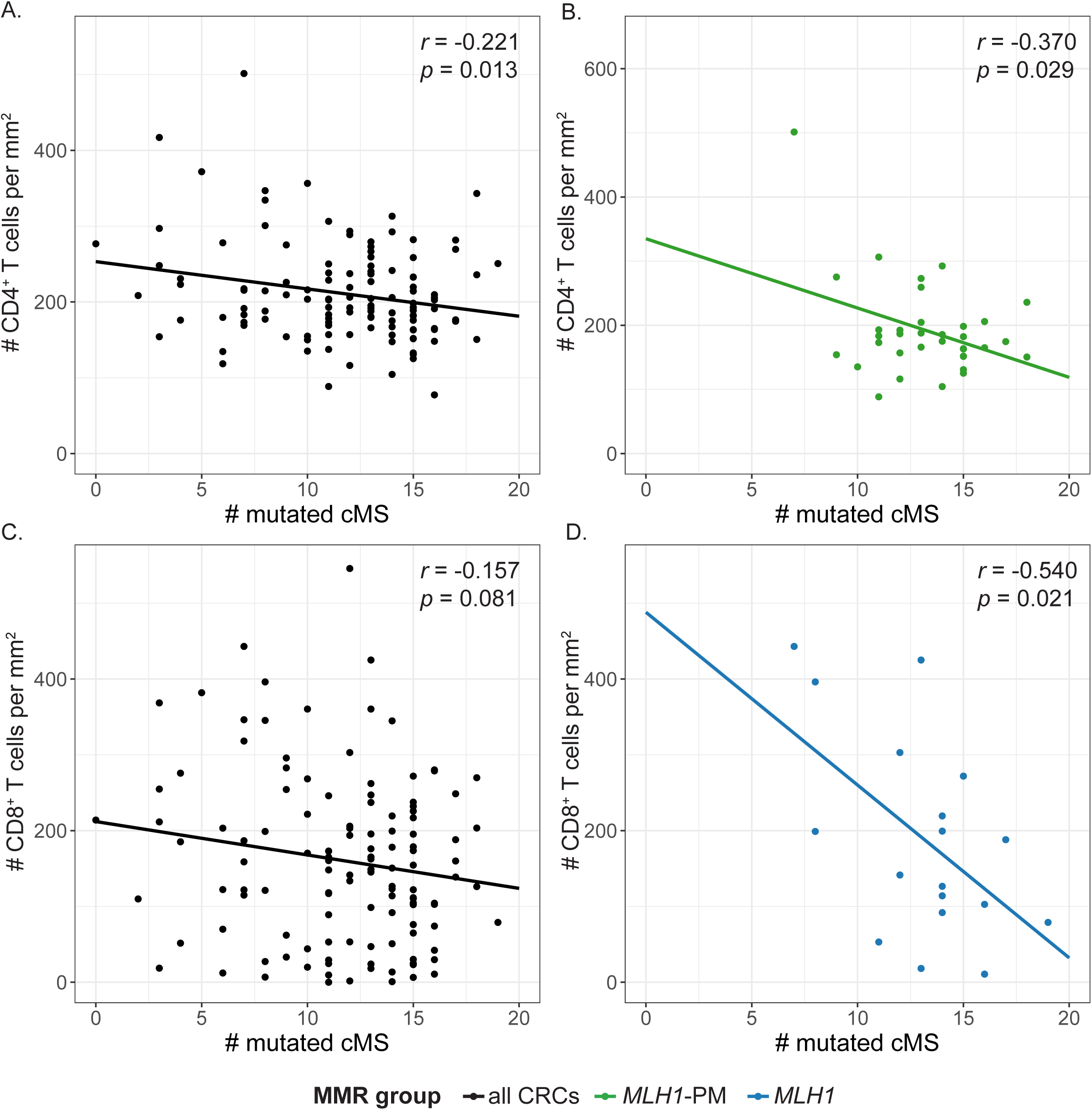
Infiltrating CD8^+^ T cell levels negatively correlate with the number of cMS mutations in *MLH1*-mutated CRCs. Correlations between the number of cMS mutations (up to 20 cMS tested per tumor) and [**A**] the number of CD4^+^ T cells in all CRCs, [**B**] CD4^+^ T cells in *MLH1*-PM CRCs [B], [**C**] CD8^+^ T cells in all CRCs, [**D**] and CD8^+^ T cells in *MLH1*-mutated CRCs. cMS mutation data was obtained from Helderman et al.^37^ *cMS, coding microsatellite; CRC, colorectal cancer; MLH1-PM, MLH1 promotor hypermethylation*.

## Discussion

Recent studies have indicated variability in both the phenotype of LS carriers^22–30^ and the genotype of LS-associated CRCs^31–37^ based on the mutated MMR gene, including potential differences in the degree of microsatellite instability (i.e. number of insertion/deletion mutations at microsatellite regions), which appears to be lowest in *MSH6*-mutated CRCs.^36, 37^ These studies have prompted the question of whether LS-associated CRCs also demonstrate diversity in immune profiles, which could potentially have implications with regard to immunotherapy management and cancer vaccine approaches and would strengthen the notion of considering LS as multiple gene-specific syndromes. However, comprehensive studies that quantitatively and qualitatively compare immune profiles of different MMR groups are scarce, applied relatively limited immunohistochemistry techniques (e.g., focusing on CD3+/CD8+ markers only) or included only a few *MSH6-* and *PMS2-*mutated CRCs.

Using IMC and mIF on one of the most extensive cohorts of LS-associated CRCs available for immunological assessment to date, our results challenge the idea that the immune profile of LS-associated CRCs depends on the specific underlying MMR defect. Instead, our findings reveal comparable immune profiles across CRCs from all MMR groups and variability within. This contrasts with our prior research, which showed lower T cell infiltration in *PMS2*-mutated CRCs compared to *MLH1*- and *MSH2*-mutated CRCs.^50^ However, the current study surpasses our earlier work in sample size and the range of included immune markers. The comparable immune response towards CRCs from every MMR group suggests that all MMR-d CRCs, independent of the MMR defect, will display sensitivity to immune checkpoint blockade. Recent evidence supporting this notion includes pathological responses observed in 100% of MMR-d CRCs compared to only 27% of MMR-proficient CRCs following neoadjuvant ipilimumab (anti-CTLA-4) plus nivolumab (anti-PD-1) treatment.^19^ However, it is noteworthy that current immunotherapy-related trials generally lack stratification of response by the MMR defect, potentially combining sporadic with hereditary tumors without specifying the underlying MMR defect.^15–19^ Consequently, a crucial step would be to validate our hypothesis by stratifying current and future immunotherapy response data based on the MMR defect.

Additional findings with potential implications for immunotherapy response include the frequent loss of HLA class I and β2M expression and the presence of γδ T cells in most tumors across all MMR groups. The increased occurrence of aberrant β2M expression in hereditary versus sporadic MMR-d CRCs with HLA class I defects aligns with our prior observations, though our earlier research did not differentiate between MMR groups.^9, 10^ This suggests that hereditary MMR-d CRCs follow distinct evolutionary pathways in tumorigenesis compared to sporadic MLH1-PM CRCs, regardless of the specific mutated MMR gene. Our current study found the prevalence of β2M defects ranging from 53% to 85%, depending on the MMR defect, which seems to exceed earlier findings of 17% to 51%.^9–11, 57–59^ Additionally, these findings do not directly align with recently published *B2M* mutation data from *MSH6*-mutated CRCs in the same cohort, where only three out of the 22 genotyped *MSH6*-mutated CRCs displayed *B2M* mutations.^37^ However, this discrepancy might be due to incomplete sequencing of the entire B2M gene in previous studies. The presence of γδ T cells, recently found to be enriched in tumors from patients with sporadic MMR-d CRCs following dual PD-1 and CTLA-4 blockade, may contribute to the immune checkpoint blockade response, particularly in patients with HLA class I-negative MMR-d CRCs, as their activation is not reliant on HLA class I binding.^56^ Our study is the first to validate their presence in hereditary MMR-d CRCs.

The observed negative correlation between the quantity of infiltrating T cells and the number of cMS mutations per tumor, particularly pronounced in *MLH1*-mutated CRCs, may imply that the acquittance of immune evasion strategies is influenced by a greater mutational burden, possibly due to heightened selective pressure. Intriguingly, since the HLA class I status remained consistent regardless of the number of cMS mutations in a tumor, mechanisms other than the loss of HLA class I expression may be at the basis of this difference, which warrant attention in forthcoming immunological studies on LS-associated CRCs.

In conclusion, by employing high-dimensional immune profiling techniques on one of the most extensive LS-associated CRC cohorts documented to date, with a comprehensive representation of *MSH6*- and *PMS2*-mutated CRCs, we illustrate that the immune profile of LS-associated CRCs remains consistent irrespective of the underlying MMR defect. This uniformity persists despite previously reported variations in the phenotype of LS carriers and the genotype of LS-associated CRCs, including varying degrees of MSI. These findings hold promise for immunotherapy strategies, suggesting sensitivity even in *MSH6-* and *PMS2-*mutated CRCs, and may contribute to a deeper understanding of the intricate interplay between tumors and the immune system in (MMR-d) CRCs and cancer in general.

## Data Availability

The datasets analyzed during this study are available from the corresponding author upon reasonable request.

## Acknowledgments

The authors sincerely thank all patients and their families for participating in this study. We gratefully acknowledge the PALGA-group collaborators/participating pathology centers for providing patient samples; T.C.T.E.F. Cronenburg for assisting in the identification of tumor regions of interest; S. van Oost for assisting in the data analysis; and R. van der Breggen for technical assistance.

## Abbreviations

β2M: β2-microglobulin
CRC: colorectal cancer
FFPE: formalin-fixed, paraffin-embedded
GZMB: granzyme B
HLA: Human Leukocyte Antigen
IHC: immunohistochemistry
IMC: imaging mass cytometry
IQR: interquartile range
LS: Lynch syndrome
mIF: multiplex immunofluorescence
*MLH1*- PM: *MLH1* promotor hypermethylation
MMR: mismatch repair
MMR-d: mismatch repair-deficiency
MSI: microsatellite instability
PD-(L)1: programmed death(-ligand) 1
SD: standard deviation.

## Figure captions

**Supplementary Figure 1. Scoring of HLA class I defects through IHC.** Illustrative examples of tumors scored positive or negative for HCA2, HC10 and β2M. *β2M, B2-microglobulin; HLA, human leukocyte antigen; IHC, immunohistochemistry*.

**Supplementary Figure 2. T cell and myeloid cell counts in the IMC dataset.** The number of [**A**] T cells, [**B**] CD4^+^ T cells, [**C**] CD8^+^ T cells, [**D**] CD103^+^ T cells, [**E**] CD103^+^ CD4^+^ T cells, [**F**] CD103^+^ CD8^+^ T cells, [**G**] granulocytes, [**H**] CD204^+^ macrophages, and [**I**] CD204^+^ CD163^+^ macrophages per tumor, stratified by MMR group. *IMC, imaging mass cytometry; MMR, mismatch repair*.

**Supplementary Figure 3. T cell condition marker expression per IMC phenotype.** Proportion of T cells positive for [**A**] CD39, [**B**] CD103, [**C**] GZMB, [**D**] ICOS, [**E**] IDO, [**F**] Ki-67, [**G**] PD-1, [**H**] Tbet, [**I**] and VISTA for each T cell phenotype per MMR group, as evaluated by IMC. Dots represent the percentage of cells per sample (calculated based on two tissue cores per sample). Black bars represent the median percentage of cells per MMR mutation group. High percentages of CD103^+^ cells in the CD103^+^ T cells, CD103^+^CD4^+^ T cells and CD103^+^CD8^+^ T cells demonstrate that our Mean-shift clustering in Cytosplore was successfully conducted. *CRC, colorectal cancer; GZMB, granzyme B; IDO, indoleamine 2,3-dioxygenase, IMC, imaging mass cytometry; MLH1-PM, MLH1 promotor hypermethylation; MMR, mismatch repair; PD-1, programmed death 1*.

**Supplementary Figure 4. Uniform myeloid cell compartments across MMR groups.** The overall number of [**A**] (HLA-DR^-/+^) CD204^+^ macrophages and [**B**] (HLA-DR^-/+^) granulocytes did not differ between the MMR groups, as evaluated by mIF. *mIF, multiplex immunofluorescence; MMR, mismatch repair*.

**Supplementary Figure 5. Correlation between the number of cMS mutations and infiltrating T cell levels per MMR group.** Correlations between the number of cMS mutations (up to 20 cMS tested per tumor) and [**A-E**] the number of CD4^+^ T cells and [**F-J**] CD4^+^ T cells per tumor, stratified by MMR group. cMS mutation data was obtained from Helderman et al.^37^ *cMS, coding microsatellite; CRC, colorectal cancer; MLH1-PM, MLH1 promotor hypermethylation*.

