## Supplementary figures and images for "Immunological profiles in Lynch syndrome colorectal cancers are not specific to mismatch repair gene defects"

### Supplementary Figure 1

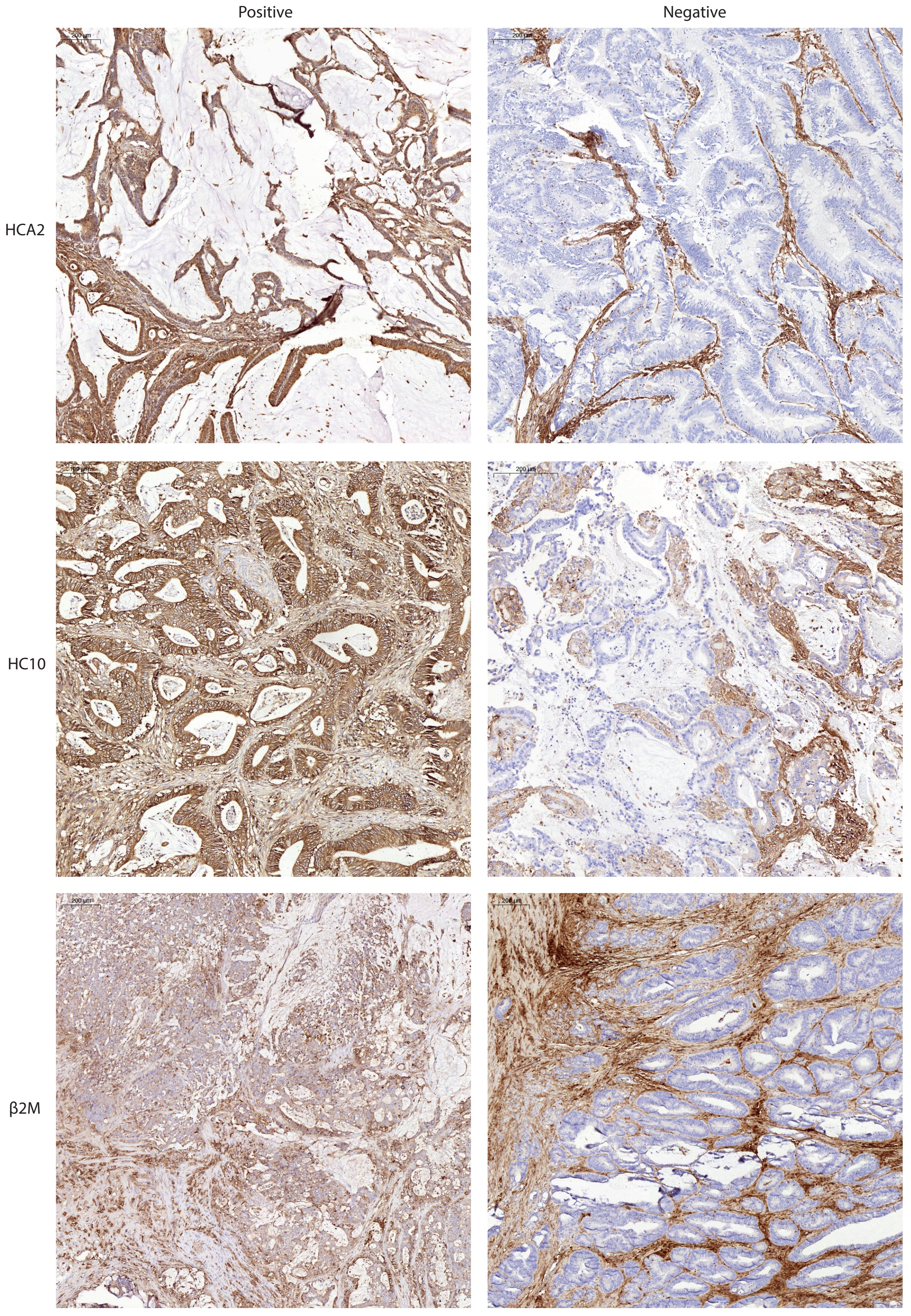

### Supplementary Figure 2

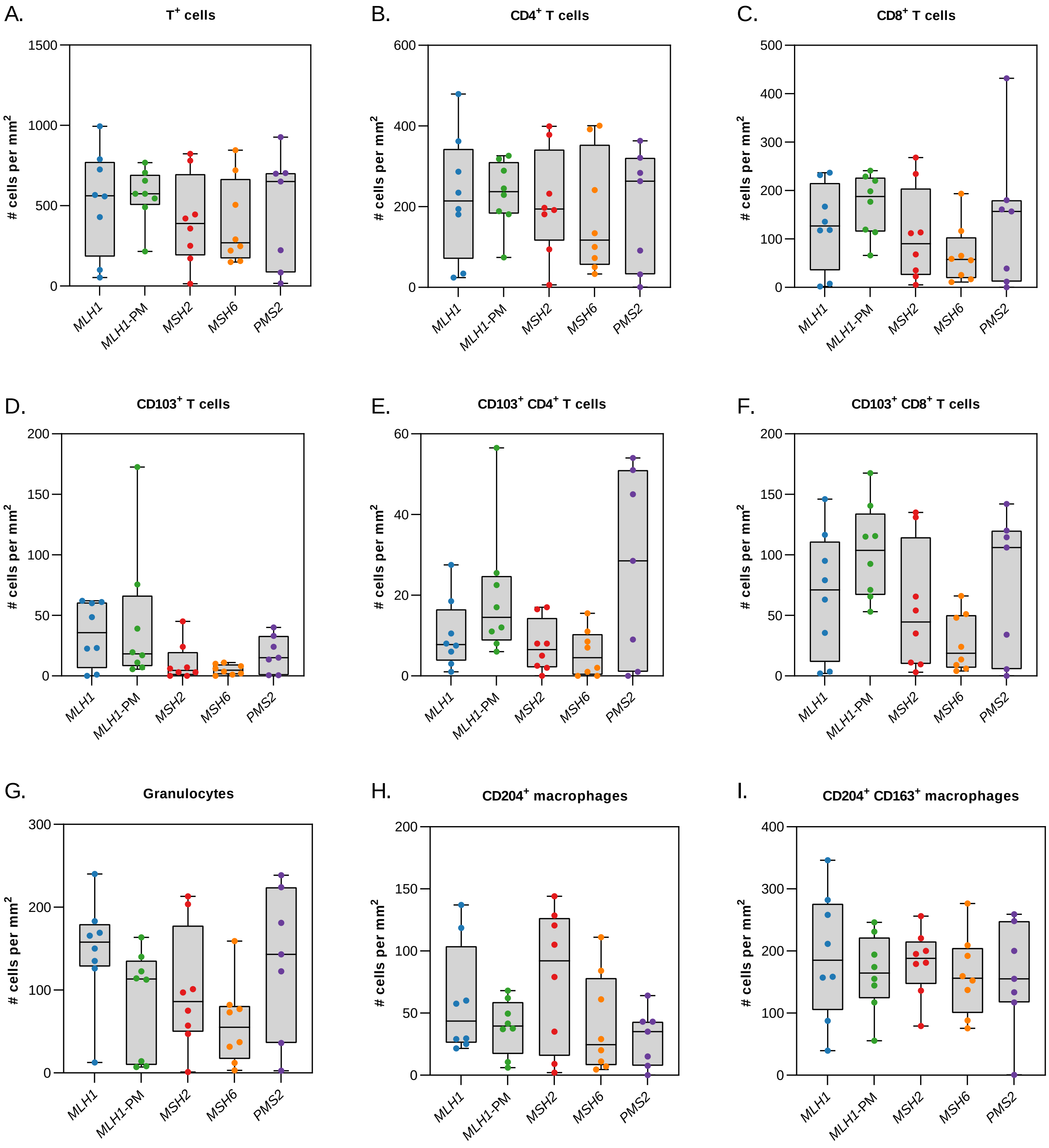

### Supplementary Figure 3

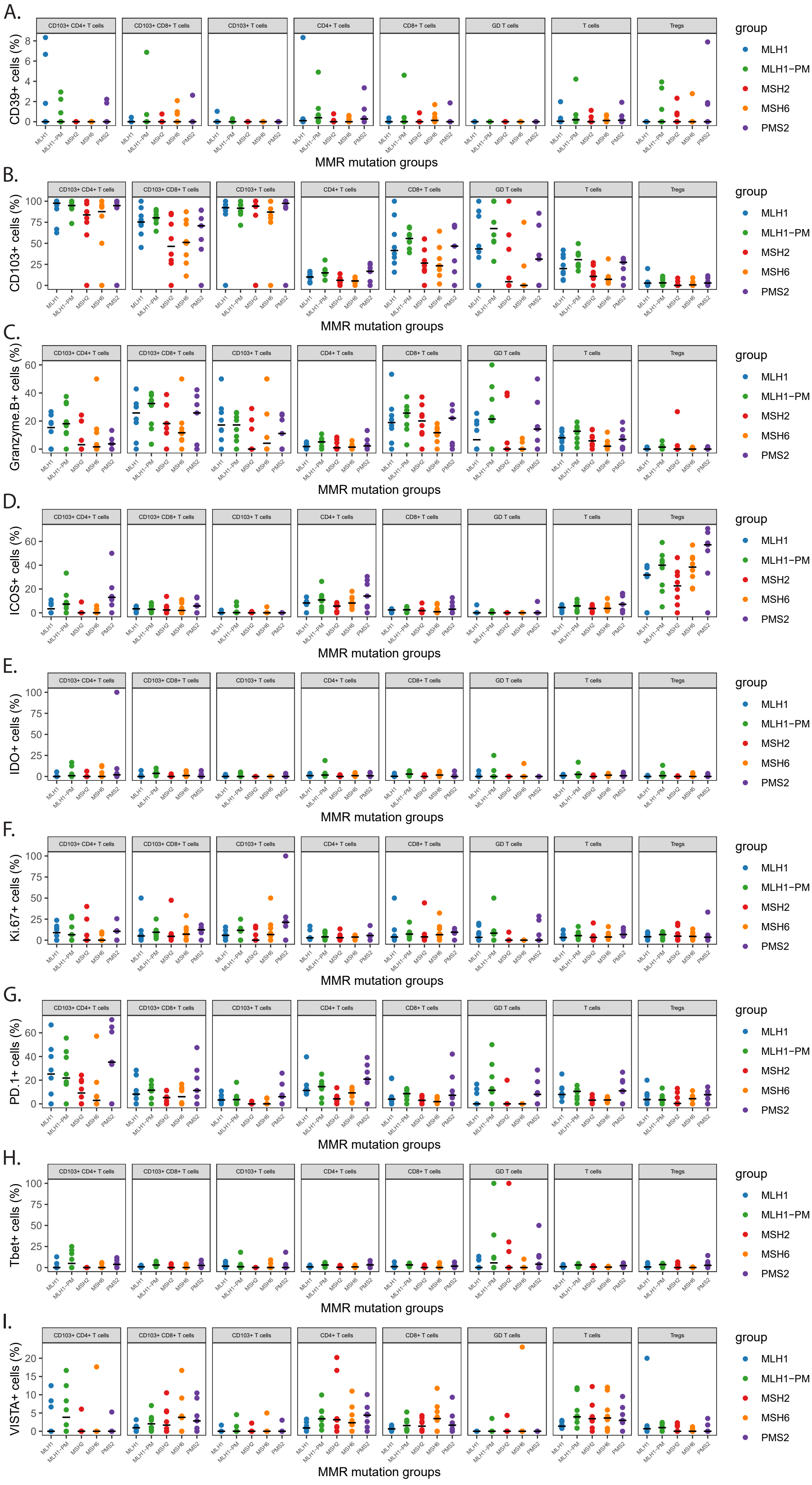

### Supplementary Figure 4

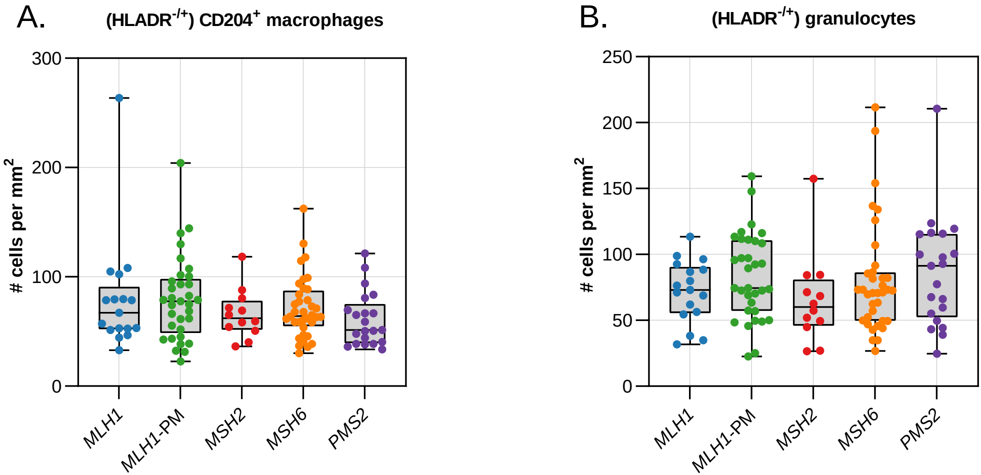

### Supplementary Figure 5

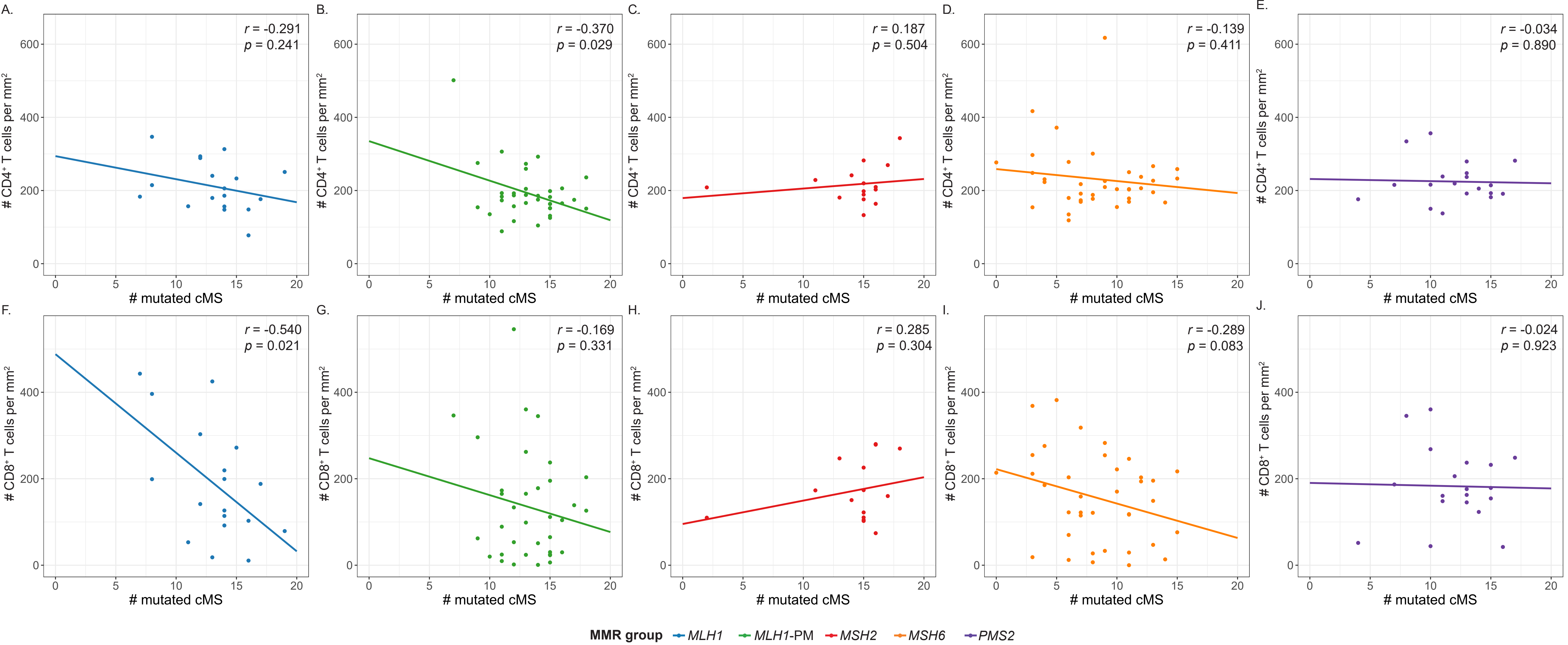
