## Supplementary Tables for "Immunological profiles in Lynch syndrome colorectal cancers are not specific to mismatch repair gene defects"

| **Supplementary Table 1. Materials and providers** | | | | | | | | | |
| --- | --- | --- | --- | --- | --- | --- | --- | --- | --- |
| **Device** | | | | | **Provider** | | | | |
| TMA Master | | | | | 3DHISTECH Kft, Budapest, Hungary | | | | |
| Hyperion mass cytometry imaging system | | | | | Fluidigm, San Francisco, CA, USA | | | | |
| 100-1000 ul Pipet-Lite XLS | | | | | Mettler-Toledo Rainin, Oakland, CA, USA | | | | |
| 20-200 ul Pipete-Lite XLS | | | | | Mettler-Toledo Rainin, Oakland, CA, USA | | | | |
| 2-20 ulPipete-Lite XLS | | | | | Mettler-Toledo Rainin, Oakland, CA, USA | | | | |
| 1-10 ulPipete-Lite XLS | | | | | Mettler-Toledo Rainin, Oakland, CA, USA | | | | |
| pH meter | | | | |  | | | | |
| Heidolph Reax top vortex mixer | | | | | Heidolph Instruments, Schwabach, Germany | | | | |
| RCA Basic Magnetic Stirrer | | | | | IKA, Staufen im Breisgau, Germany | | | | |
| Microwave | | | | | Panasonic, Kadoma, Osaka, Japan | | | | |
| Zeiss Axioskop 20 451497 | | | | | Oberkochen, Baden-Wurttemberg, Germany | | | | |
| Vectra® 3 Automated Quantitative Pathology Imaging System | | | | | Akoya Biosciences, Marlborough, MA, United States | | | | |
| **Chemical/reagents** | | | | | **Provider** | | | | |
| 0.3% hydrogen/peroxidase methanol solution | | | | | Merck Millipore, Burlington, MA, USA | | | | |
| BrightVision 1 step detection system Goat Anti-Mouse/Rabbit HRP | | | | | Immunologic, Duiven, The Netherlands | | | | |
| DAB+ chromogen | | | | | DAKO, Agilent technologies, Santa Clara, CA, USA | | | | |
| Hematoxylin | | | | | Thermo Fisher Scientific, Waltham, MA, USA | | | | |
| Hematoxylin er (Mayer) | | | | | Avantor Performance Materials Poland, Gliwice, Poland | | | | |
| Antibody stabilizer solution | | | | | Candor Bioscience, Wangen im Allgäu, Germany | | | | |
| W-buffer | | | | | Fluidigm, San Francisco, CA, USA | | | | |
| Superblock solution | | | | | Thermo Fisher Scientific, Waltham, MA, USA | | | | |
| EMSURE® ACS, ISO, Reag. Ph Eur Methanol for analysis | | | | | Merck Millipore, Burlington, MA, USA | | | | |
| 1X Plus Amplification Diluent | | | | | PerkinElmer, Waltham, MA, USA | | | | |
| PBS/BSA 1% | | | | | Sigma Aldrich, MO, USA | | | | |
| ProLong® Gold Antifade Reagent | | | | | Cell signaling Technologies, MA, USA | | | | |
| Surgipath Micromount Mounting Medium | | | | | Leica Biosystems Richmond, Ruchmond, IL, USA | | | | |
| Xylol | | | | | Merck Millipore, Burlington, MA, USA | | | | |
| **Consumable** | | | | | **Provider** | | | | |
| Silane-coated glass slides | | | | | VWR, Radnor, PA, USA | | | | |
| Eppendorf Safe-Lock Tube 1.5 ml | | | | | Eppendorf SE, Hamburg, Germany | | | | |
| Eppendorf Safe-Lock Tube 2 ml | | | | | Eppendorf SE, Hamburg, Germany | | | | |
| Epredia Cover Slips 24x40 mm | | | | | Gerhard Menzel Gmbh, Braunschweig, Germany | | | | |
| PE Pasteur Pipette 1 ml | | | | | LP ITALIANA SPA, Milan, Italy | | | | |
| Pipette Tips GPS LTS 1000µL 768A/8 | | | | | Mettler-Toledo Rainin, Oakland, CA, USA | | | | |
| Pipette Tips GPS LTS 20µL 960A/10 | | | | | Mettler-Toledo Rainin, Oakland, CA, USA | | | | |
| Pipette Tips GPS LTS 250µL 960A/10 | | | | | Mettler-Toledo Rainin, Oakland, CA, USA | | | | |
| Tube, 15 ml, PP, 17/120 mm, conical bottom, blue screw cap, steril | | | | | Greiner Bio-One, Alphen aan den Rijn, The Netherlands | | | | |
| Tube, 50 ml, PP, graduated, conical bottom, blue screw cap, steril | | | | | Greiner Bio-One, Alphen aan den Rijn, TheNetherlands | | | | |
| **Kits** | | | | | **Provider** | | | | |
| Maxpar antibody labeling kit | | | | | Fluidigm, San Francisco, CA, USA | | | | |
| **Antibody** | **Technique** | **Clone** | | **Metal** | **Incubation time** | **Incubation temp** | | **Dilution** | **Provider** |
| HLA class I^a^ | IHC | HCA2 | | NA | Indirect ON | 4C | | 1000 | nordic-mubio |
| HLA class I^b^ | IHC | HC10 | | NA | Indirect ON | 4C | | 3200 | nordic-mubio |
| β2M | IHC | EPR21752-214 | | NA | Indirect ON | 4C | | 4000 | abcam |
| PD-L1 | IHC | E1L3N | | NA | Indirect ON | 4C | | 200 | cst |
| TCRδ | IHC | H-41 | | NA | Indirect ON | 4C | | 200 | santa cruz |
| CD8a | IMC | D8A8Y | | 146 Nd | 5h | RT | | 50 | cst |
| PD-1 | IMC | D4W2J | | 160 Gd | 5h | RT | | 50 | cst |
| ICOS | IMC | D1K2T(tm) | | 161 Dy | 5h | RT | | 50 | cst |
| CD204 | IMC | J5HTR3 | | 164 Dy | 5h | RT | | 50 | thermo |
| CD103 | IMC | EPR4166(2) | | 168 Er | 5h | RT | | 50 | abcam |
| Tbet | IMC | 4B10 | | 170 Er | 5h | RT | | 50 | biolegend |
| CD19 | IMC | D4V4B | | 172 Yb | 5h | RT | | 50 | cst |
| CD163 | IMC | D6U1J | | 173 Yb | 5h | RT | | 50 | cst |
| TGFbeta | IMC | TB21 | | 115In | 5h | RT | | 100 | thermo |
| HLA-DR | IMC | TAL 1B5 | | 141 Pr | 5h | RT | | 100 | abcam |
| CD11b | IMC | D6X1N | | 144 Nd | 5h | RT | | 100 | cst |
| Granzyme B | IMC | D6E9W | | 150 Nd | 5h | RT | | 100 | cst |
| cleaved caspase | IMC | 5A1E | | 155 Gd | 5h | RT | | 100 | cst |
| CD39 | IMC | EPR20627 | | 157 Gd | 5h | RT | | 100 | abcam |
| VISTA | IMC | D1L2G(TM) | | 158 Gd | 5h | RT | | 100 | cst |
| CD14 | IMC | D7A2T | | 163 Dy | 5h | RT | | 100 | cst |
| CD56 | IMC | E7X9M | | 167 Er | 5h | RT | | 100 | cst |
| CD7 | IMC | EPR4242 | | 174 Yb | 5h | RT | | 100 | abcam |
| CD11c | IMC | EP1347Y | | 176 Yb | 5h | RT | | 100 | abcam |
| TCRgd + 2nd Ab | IMC | H41 | | 148 Nd | Indirect ON | 4C | | 50 | santa cruz |
| CD4 +2nd AB | IMC | EPR6855 | | 145 Nd | Indirect ON | RT | | 100 | abcam |
| CD45 | IMC | D9M8I | | 149 Sm | ON | 4C | | 50 | cst |
| CD3 | IMC | EP449E | | 153 Eu | ON | 4C | | 50 | abcam |
| PD-L1 | IMC | E1L3N(R) | | 156 Gd | ON | 4C | | 50 | cst |
| FOXP3 | IMC | D608R | | 159 Tb | ON | 4C | | 50 | cst |
| CD27 | IMC | EPR8569 | | 175 Lu | ON | 4C | | 50 | abcam |
| Vimentin | IMC | D21H3 | | 194 Pt | ON | 4C | | 50 | cst |
| Keratin | IMC | C11 and AE1/AE3 | | 198 Pt | ON | 4C | | 50 | biolegend/cst |
| B catenin | IMC | D10A8 | | 89Y | ON | 4C | | 100 | cst |
| CD20 | IMC | H1 | | 142 Nd | ON | 4C | | 100 | BD |
| CD68 | IMC | D4B9C | | 143 Nd | ON | 4C | | 100 | cst |
| CD31 | IMC | 89C2 | | 147 Sm | ON | 4C | | 100 | cst |
| CD57 | IMC | HNK-1 / Leu-7 | | 151 Eu | ON | 4C | | 100 | abcam |
| Ki-67 | IMC | 8D5 | | 152 Sm | ON | 4C | | 100 | cst |
| P16ink4a | IMC | D3W8G | | 154 Sm | ON | 4C | | 100 | cst |
| IDO | IMC | D5J4E(TM) | | 162 Dy | ON | 4C | | 100 | cst |
| CD45RO | IMC | UCHL1 | | 165 Ho | ON | 4C | | 100 | cst |
| D2-40 | IMC | D2-40 | | 166 Er | ON | 4C | | 100 | biolegend |
| CD38 | IMC | EPR4106 | | 169 Tm | ON | 4C | | 100 | abcam |
| CD15 | IMC | MC480 | | 171 Yb | ON | 4C | | 100 | cst |
| Histone H3 | IMC | D1H2 | | 209 | ON | 4C | | 50 | cst |
| CD4 | mIF | EPR6855 | | NA | Indirect 1h | RT | | 1000 | abcam |
| CD8a | mIF | D8A8Y | | NA | Indirect 1h | RT | | 250 | cst |
| CD103 | mIF | EPR4166(2) | | NA | Indirect 1h | RT | | 1000 | abcam |
| GZMB | mIF | D6E9W | | NA | Indirect 1h | RT | | 125 | cst |
| PD-1 | mIF | D4W2J | | NA | Indirect 1h | RT | | 125 | cst |
| CD204 | mIF | J5HTR3 | | NA | Indirect 1h | RT | | 1000 | thermo |
| HLA-DR | mIF | TAL 1B5 | | NA | Indirect 1h | RT | | 1000 | thermo |
| CD15/SSEA1 | mIF | MC480 | | NA | Indirect 1h | RT | | 1000 | cst |
| **Opals** | | | | | **Provider** | | | | |
| Opal 520 (FP1487A) | | | | | Akoya Biosciences, Marlborough, MA, USA | | | | |
| Opal 570 (FP1488A) | | | | | Akoya Biosciences, Marlborough, MA, USA | | | | |
| Opal 620 (FP1495) | | | | | Akoya Biosciences, Marlborough, MA, USA | | | | |
| Opal 650 (FP1496) | | | | | Akoya Biosciences, Marlborough, MA, USA | | | | |
| Opal 690 (FP1497) | | | | | Akoya Biosciences, Marlborough, MA, USA | | | | |
| **Software** | | | **Description** | | | | **Provider** | | |
| R v4.1.3 | | | Statistical analysis | | | | R Foundation for Statistical Computing, Vienna, Austria | | |
| RStudio v2022.02.3+492 | | | Statistical analysis | | | | RStudio Team, Boston, MA, USA | | |
| IBM SPSS Statistics v29.0 | | | Statistical analysis | | | | International Business Machines Corporation (IBM), NY, USA | | |
| CaseViewer v2.4.0 | | | Visualization of IHC scans | | | | 3DHISTECH Ltd., Budapest, Hungary | | |
| MCD viewer 1.0.560.6 | | | Visualization of MCD files | | | | Fluidigm, San Francisco, CA, USA | | |
| MATLAB R2021b | | | Image enhancement (IMC) | | | | MathWorks, Natick, MA, USA | | |
| Ilastik v.1.4.0 | | | Semi-automated background-removal (IMC) | | | | <https://www.ilastik.org/download> | | |
| CellProfiler v4.2.1/v.4.2.4 | | | Creation of single cell masks (IMC), cell segmentation (mIF) | | | | <https://cellprofiler.org/releases> | | |
| ImaCytE | | | Single cell clustering (IMC), interaction analysis (IMC) | | | | <https://github.com/biovault/ImaCytE.git> | | |
| QuPath v0.4.3 | | | Analysis of IHC/mIF data | | | | QuPath developers, The University of Edinburgh | | |
| Phenochart Software | | | Scanning of mIF slides | | | | Akoya Biosciences, Marlborough, MA, United States | | |
| inForm® analysis software | | | Spectral separation of dyes for mIF analysis | | | | Akoya Biosciences, Marlborough, MA, United States | | |
| PENGUIN | | | Image normalization and rescaling (mIF) | | | |  | | |
| FlowSOM v.2.6.0 | | | Cluster cells by mean marker intensity (mIF) | | | | <https://bioconductor.org/packages/FlowSOM/> | | |
| ^a^β2M-free HLA-A (except -A24), -B7301 and -G heavy chains  ^b^β2M-free HLA-B and C heavy chains and on β2m-free HLA-A10, -A28, -A29, -A30, -A31, -A32 and -A33 heavy chains  *IHC, immunohistochemistry; IMC, imaging mass cytometry; mIF, immunofluorescence; NA, not applicable/not available; ON, overnight; RT, room temperature* | | | | | | | | | |

| **Supplementary Table 2. Expression of markers corresponding to each identified phenotype in the** **IMC dataset** | |
| --- | --- |
| **Phenotype** | **Marker expression^a^** |
| Apoptotic cancer cells | Keratin/B catenin, cleaved caspase |
| B cells | CD20 |
| CD103+ CD4+ T cells | CD3, CD45, CD4, CD103 |
| CD103+ CD8+ T cells | CD3, CD45, CD8a, CD103 |
| CD103+ T cells | CD3, CD45, CD103 |
| CD11b+ cells | CD11b |
| CD204+ CD163+ macrophages | CD68, CD204, CD163 |
| CD204+ macrophages | CD68, CD204 |
| CD4+ T cells | CD3, CD45, CD4 |
| CD8+ T cells | CD3, CD45, CD8a |
| D2-40 fibroblast | Vimentin, D2-40 |
| Dendritic cells | CD11c |
| Fibroblast | Vimentin |
| GD T cells | CD3, CD45, GD |
| Granulocyte | CD11b, CD15 |
| IDO+ cancer cells | Keratin/B catenin, IDO |
| Innate lymphoid cells | CD3, CD45, CD7 |
| Lymph vessel | CD31, D2-40 |
| Monocytes | CD14 |
| Other macrophages | CD68, CD11c |
| Plasma cells/plasmablasts | CD38 |
| Proliferating cancer cells | Keratin/B catenin, Ki-67 |
| p16^+^ cancer cells | Keratin/B catenin, p16ink4a |
| T cells | CD3, CD45 |
| Tregs | CD3, CD45, FOXP3 |
| Cancer cells | Keratin/B catenin |
| Vessel | CD31 |
| ^a^The markers presented were crucial criteria for classifying cells into their designated phenotypes. It is possible that additional markers, not explicitly mentioned here, also have been expressed. | |

| **Supplementary Table 3. Cohort description** | | | | | | |
| --- | --- | --- | --- | --- | --- | --- |
| **Description** | | ***MLH1* (n=18)** | ***MLH1*-PM (n=35)** | ***MSH2* (n=16)** | ***MSH6* (n=40)** | ***PMS2* (n=23)** |
| **Sex [male], n (%)** | | 9 (50%) | 6 (17.1) | 11 (68.8) | 24 (60%) | 11 (47.8) |
| **Age at CRC diagnosis [years]** | |  |  |  |  |  |
|  | Median (IQR) | 48 (41-60.3) | 66 (59-71) | 51 (38.5-58.75) | 65 (47-71) | 46 (41-58) |
|  | Mean (SD) | 50.1 (12.4) | 65.94 (9.4) | 48.25 (13) | 61.7 (13.8) | 49.1 (11.1) |
|  | NA | 0 | 0 | 0 | 1 | 0 |
| **Side, n (%)** | |  |  |  |  |  |
|  | Right^a^ | 13 (72.2) | 32 (91.4) | 11 (68.8) | 19 (47.5) | 14 (60.9) |
|  | Left^b^ | 2 (11.1) | 2 (5.7) | 5 (31.3) | 19 (47.5) | 9 (39.1) |
|  | NA | 3 (16.7) | 1 (2.9) | 0 | 2 (5) |  |
| **Location, n (%)** | |  |  |  |  |  |
|  | Cecum/ascending colon | 11 (61.1) | 19 (54.3) | 8 (50) | 13 (32.5) | 10 (43.5) |
|  | Transverse colon | 2 (11.1) | 12 (34.3) | 3 (18.8) | 4 (10) | 1 (4.3) |
|  | Descending colon | 1 (5.6) | 1 (2.9) | 1 (6.3) | 3 (7.5) | 1 (4.3) |
|  | Rectosigmoid | 1 (5.6) | 1 (2.9) | 4 (25) | 16 (40) | 8 (34.8) |
|  | NA | 3 (16.7) | 2 (5.7) | 0 | 4 (10) | 3 (13) |
| **Differentiation, n (%)** | |  |  |  |  |  |
|  | Well | 0 | 0 | 1 (6.3) | 3 (7.5) | 1 (4.3) |
|  | Well/moderate | 4 (22.2) | 6 (17.1) | 4 (25) | 5 (12.5) | 6 (26.1) |
|  | Moderate | 7 (38.9) | 7 (20) | 7 (43.8) | 19 (47.5) | 9 (39.1) |
|  | Moderate/poor | 1 (5.6) | 6 (17.1) | 2 (12.5) | 4 (10) | 2 (8.7) |
|  | Poor | 1 (5.6) | 7 (20) | 0 | 5 (12.5) | 2 (8.7) |
|  | NA | 5 (27.8) | 9 (25.7) | 2 (12.5) | 4 (10) | 3 (13) |
| **Diameter [cm]** | |  |  |  |  |  |
|  | Median (IQR) | 3.8 (2-5) | 4.9 (2.6-7) | 2.5 (2-6) | 5.5 (3.1-7) | 5 (3.5-6.3) |
|  | Mean (SD) | 5.2 (5.8) | 5.0 (2.4) | 5.5 (7) | 6.2 (7.4) | 6.6 (7.4) |
|  | NA | 3 | 3 | 5 | 8 | 5 |
| **MSI status, n (%)** | |  |  |  |  |  |
|  | MSS | 0 | 0 | 1 (6.3) | 2 (5) | 3 (13) |
|  | MSI | 18 (100) | 35 (100) | 15 (93.8) | 37 (92.5) | 20 (87) |
|  | NA | 0 | 0 | 0 | 1 | 0 |
| **IHC status, n (%)^c^** | |  |  |  |  |  |
|  | Loss of MMR expression | 18 (100) | 35 (100) | 16 (100) | 30 (75) | 6 (26) |
|  | Normal MMR expression | 0 | 0 | 0 | 3 (7.5) | 1 (4) |
|  | NA | 0 | 0 | 0 | 7 (18) | 16 (70) |
| ^a^Right-sided locations include the cecum/ascending colon and transverse colon.  ^b^Left-sided locations include the descending colon, sigmoid colon and rectum.  ^c^Only the loss of expression of the MMR protein that corresponds with the constitutional MMR variant is taken into account.  *CRC, colorectal cancer; IHC, immunohistochemistry; IQR, interquartile range; MMR, mismatch repair; MLH1-PM, MLH1 promotor hypermethylation; MSI, microsatellite instability; MSS, microsatellite stable; NA, not applicable/not available;* | | | | | | |

| **Supplementary Table 4. Clinical and histological characteristics of all analyzed CRCs** | | | | | | | | | | | | |
| --- | --- | --- | --- | --- | --- | --- | --- | --- | --- | --- | --- | --- |
| **Study ID** | **Sex** | **Age at CRC diagnosis [5-year range]** | **Tumor side** | **Tumor location** | **Differentiation** | **Diameter [cm]** | **MSI** | **IHC** | | | | |
|  |  |  |  |  |  |  |  | **MMR** | **HCA2 defect** | **HC10 defect** | **B2M defect** | **PD-L1 expression** |
| MLH1_01 | f | 71-75 | right | cecum/ascending colon | moderate/well | 1.5 | MSI | MLH1- | y | n | y | n |
| MLH1_02 | f | 61-65 |  |  |  | 3.8 | MSI | MLH1- | n | n | n | n |
| MLH1_03 | f | 51-55 | right | transverse colon | moderate | 1.2 | MSI | MLH1- | y | n | n | n |
| MLH1_04 | m | 51-55 | right | cecum/ascending colon | moderate | 2 | MSI | MLH1- | y | n | n | n |
| MLH1_05 | m | 41-45 | left | descending colon | moderate/well | 11 | MSI | MLH1- | y | y | y | n |
| MLH1_06 | f | 61-65 | right | cecum/ascending colon | moderate | 4 | MSI | MLH1- | y | y | y | n |
| MLH1_07 | m | 41-45 | right | cecum/ascending colon |  | 5 | MSI | MLH1-MSH2+MSH6+PMS2- | y | y | y | n |
| MLH1_08 | f | 41-45 | right | cecum/ascending colon |  | 2.5 | MSI | MLH1- | n | n | y | n |
| MLH1_09 | f | 31-35 | right | cecum/ascending colon | moderate | 1 | MSI | MLH1- | y | y | y | n |
| MLH1_10 | m | 36-40 | right | cecum/ascending colon | moderate | 3.5 | MSI | MLH1- | n | n | y | n |
| MLH1_11 | m | 41-45 | right | cecum/ascending colon |  | 3 | MSI | MLH1- | y | n | y | n |
| MLH1_12 | f | 31-35 | right | cecum/ascending colon | moderate |  | MSI | MLH1- | n | n | n | n |
| MLH1_13 | m | 41-45 | right | transverse colon | poor | 4 | MSI | MLH1-MSH2+MSH6+PMS2+ | y | y | y | n |
| MLH1_14 | f | 76-80 | right | cecum/ascending colon | moderate/well | 2.4 | MSI | MLH1- | y | y | y | n |
| MLH1_15 | m | 46-50 | right | cecum/ascending colon | poor/moderate |  | MSI | MLH1-MSH2+MSH6+PMS2- | y | n | y | n |
| MLH1_16 | m | 51-55 | left | rectosigmoid | moderate/well | 5 | MSI | MLH1- | y | y | y | n |
| MLH1_17 | f | 56-60 |  |  | moderate | 7 | MSI | MLH1- | y | y | y | n |
| MLH1_18 | m | 46-50 |  |  |  |  | MSI | MLH1-MSH2+MSH6+PMS2- | n | n | y | n |
| MLH1-PM_01 | f | 91-95 | right | cecum/ascending colon | poor/moderate | 8.5 | MSI | MLH1-MSH2+MSH6-PMS2- | y | y | n | y |
| MLH1-PM_02 | f | 66-70 | right | cecum/ascending colon | poor/moderate | 2.5 | MSI | MLH1-MSH2+MSH6+PMS2- | y | y | n | y |
| MLH1-PM_03 | f | 66-70 | right | transverse colon | moderate/well | 2.3 | MSI | MLH1-MSH2+MSH6+PMS2- | n | y | n | n |
| MLH1-PM_04 | f | 66-70 | right | transverse colon |  | 2.5 | MSI | MLH1-MSH2+MSH6+PMS2- | y | y | y | n |
| MLH1-PM_05 | f | 71-75 | right | transverse colon | moderate/well | 4.3 | MSI | MLH1-MSH2+MSH6+PMS2- | y | y | y | n |
| MLH1-PM_06 | m | 61-65 | right | cecum/ascending colon | poor/moderate | 8 | MSI | MLH1-MSH2+MSH6+PMS2- | y | y | n | n |
| MLH1-PM_07 | m | 66-70 | right | transverse colon | moderate/well | 1.8 | MSI | MLH1-MSH2+MSH6+PMS2- | y | y | n | n |
| MLH1-PM_08 | f | 61-65 | right | transverse colon | moderate/well | 4 | MSI | MLH1-MSH2+MSH6+PMS2- | y | y | n | n |
| MLH1-PM_09 | m | 66-70 | right |  | poor/moderate | 1 | MSI | MLH1-MSH2+MSH6+PMS2- | n | n |  | n |
| MLH1-PM_10 | m | 51-55 | right | cecum/ascending colon | moderate/well | 1.5 | MSI | MLH1-MSH2+MSH6+PMS2- | y | y | y | n |
| MLH1-PM_11 | f | 76-80 | right | cecum/ascending colon | poor/moderate | 3.5 | MSI | MLH1-MSH2+MSH6+PMS2- | n | y | y | y |
| MLH1-PM_12 | f | 71-75 | right | cecum/ascending colon | poor | 5 | MSI | MLH1-MSH2+MSH6+PMS2- | y | y |  | n |
| MLH1-PM_13 | f | 71-75 | right | transverse colon | poor | 5 | MSI | MLH1- | y | n |  | y |
| MLH1-PM_14 | f | 66-70 | right | cecum/ascending colon |  | 2 | MSI | MLH1-MSH2+MSH6+PMS2- | y | y | n | n |
| MLH1-PM_15 | f | 66-70 | right | transverse colon |  | 5.5 | MSI | MLH1-MSH2+MSH6+PMS2- | n | y | n | n |
| MLH1-PM_16 | f | 61-65 | left | rectosigmoid | poor | 8.5 | MSI | MLH1-MSH2+MSH6+PMS2- |  | n | n | n |
| MLH1-PM_17 | f | 66-70 | right | transverse colon | moderate | 7 | MSI | MLH1-MSH2+MSH6+PMS2- | y | y | n | n |
| MLH1-PM_18 | f | 71-75 | right | transverse colon | poor | 9 | MSI | MLH1- | y | y | n | n |
| MLH1-PM_19 | f | 51-55 | right | cecum/ascending colon | moderate | 7 | MSI | MLH1-MSH2+MSH6+PMS2- | y | y | y | n |
| MLH1-PM_20 | f | 61-65 | right | cecum/ascending colon | poor | 4.5 | MSI | MLH1- | y | y | y | y |
| MLH1-PM_21 | f | 71-75 |  |  | poor |  | MSI | MLH1-MSH2+MSH6+PMS2- | y | y | y | n |
| MLH1-PM_22 | f | 76-80 | right | cecum/ascending colon |  | 8 | MSI | MLH1- | n | n | n | n |
| MLH1-PM_23 | f | 61-65 | left | descending colon | poor | 9.5 | MSI | MLH1-MSH2+MSH6+PMS2- | y | y | n |  |
| MLH1-PM_24 | f | 51-55 | right | cecum/ascending colon | moderate | 5.2 | MSI | MLH1-MSH2+MSH6+PMS2- | y | y | y | n |
| MLH1-PM_25 | f | 56-60 | right | cecum/ascending colon | poor/moderate | 4.5 | MSI | MLH1-MSH2+MSH6+PMS2- | y | y | n | n |
| MLH1-PM_26 | f | 46-50 | right | cecum/ascending colon | moderate | 2.5 | MSI | MLH1- | y | y | n | y |
| MLH1-PM_27 | f | 56-60 | right | cecum/ascending colon |  | 6 | MSI | MLH1-MSH2+MSH6+PMS2- | n | n | y | n |
| MLH1-PM_28 | m | 56-60 | right | cecum/ascending colon |  | 8 | MSI | MLH1-MSH2+MSH6+PMS2- | y | y | n | n |
| MLH1-PM_29 | f | 56-60 | right | transverse colon | moderate |  | MSI | MLH1-MSH2+MSH6+PMS2- | n | n | n | n |
| MLH1-PM_30 | f | 51-55 | right | transverse colon |  | 5 | MSI | MLH1-MSH2+MSH6+PMS2- | n | y | y | n |
| MLH1-PM_31 | f | 71-75 | right | cecum/ascending colon | moderate | 4.2 | MSI | MLH1-MSH2+MSH6+PMS2- | n | y | y | n |
| MLH1-PM_32 | f | 66-70 | right | cecum/ascending colon |  |  | MSI | MLH1-MSH2+MSH6+PMS2- | y | y | n | n |
| MLH1-PM_33 | f | 61-65 | right | cecum/ascending colon |  | 4.7 | MSI | MLH1-MSH2+MSH6+PMS2- | n | n | n | n |
| MLH1-PM_34 | f | 56-60 | right | cecum/ascending colon | moderate | 6 | MSI | MLH1-MSH2+MSH6+PMS2- | n | n | n | n |
| MLH1-PM_35 | m | 71-75 | right | transverse colon | moderate/well | 2.9 | MSI | MLH1-MSH2+MSH6+PMS2- | n | n | n | n |
| MSH2_01 | f | 56-60 | left | rectosigmoid | moderate | 1.5 | MSI | MSH2- | y | y | y | n |
| MSH2_02 | m | 46-50 | right | transverse colon | moderate/well | 10 | MSI | MLH1+MSH2-MSH6-PMS2+ | n | n | n | n |
| MSH2_03 | m | 56-60 | left | rectosigmoid | moderate |  | MSI | MSH2- | y | y | y | n |
| MSH2_04 | m | 26-30 | right | cecum/ascending colon | moderate |  | MSI | MSH2- | y | y | y | n |
| MSH2_05 | m | 46-50 | right | transverse colon | moderate/well | 2 | MSI | MLH1+MSH2-MSH6-PMS2+ | y | n | n | n |
| MSH2_06 | m | 36-40 | right | cecum/ascending colon | well | 3.7 | MSI | MLH1-MSH2-MSH6+PMS2- | y | y | y | n |
| MSH2_07 | m | 26-30 | left | rectosigmoid |  | 5 | MSI | MLH1+MSH2-MSH6-PMS2+ | y | n | n | y |
| MSH2_08 | m | 61-65 | right | cecum/ascending colon | moderate/well | 0.3 | MSS | MLH1+MSH2-MSH6-/+PMS2+ | n | n | n | n |
| MSH2_09 | m | 61-65 | right | cecum/ascending colon |  | 2 | MSI | MLH1+MSH2-MSH6-PMS2+ | n | n | n |  |
| MSH2_10 | f | 41-45 | right | transverse colon | moderate | 2.2 | MSI | MLH1+MSH2-MSH6+PMS2- | y | y | y | n |
| MSH2_11 | f | 26-30 | left | rectosigmoid | poor/moderate |  | MSI | MLH1+MSH2-MSH6-PMS2+ | y |  | n | n |
| MSH2_12 | m | 51-55 | right | cecum/ascending colon | poor/moderate | 6 | MSI | MLH1+MSH2-MSH6-PMS2+ | y | y | y | y |
| MSH2_13 | m | 51-55 | left | descending colon | moderate/well |  | MSI | MLH1+MSH2-MSH6- | y | y | n | n |
| MSH2_14 | m | 36-40 | right | cecum/ascending colon | moderate | 2.5 | MSI | MLH1+MSH2-MSH6-PMS2+ | n | y | n | n |
| MSH2_15 | f | 56-60 | right | cecum/ascending colon | moderate |  | MSI | MSH2- | n | n | y | n |
| MSH2_16 | f | 61-65 | right | cecum/ascending colon | moderate | 2.5 | MSI | MLH1+MSH2-MSH6-PMS2+ |  |  | y |  |
| MSH6_01 | f | 76-80 | right | cecum/ascending colon | moderate | 4 | MSI | MSH6- | y | n | n | n |
| MSH6_02 | m | 66-70 | right | cecum/ascending colon | well | 5.5 | MSI | MLH1+MSH2-MSH6-PMS2+ | y | y | n | n |
| MSH6_03 | f | 81-85 | left | rectosigmoid | poor |  |  |  | y | y | y | n |
| MSH6_04 | f | 46-50 | left | descending colon | poor/moderate | 7.5 | MSI | MLH1+MSH2+MSH6+PMS2+ | y | y | y | y |
| MSH6_05 | m | 56-60 | right | transverse colon | moderate | 5.4 | MSI | MLH1+MSH2+MSH6-/+PMS2+ | y | y | y | n |
| MSH6_06 | m | 66-70 | right | cecum/ascending colon | well | 0.7 | MSI | MLH1+MSH2+MSH6-PMS2+ | y | y | n | n |
| MSH6_07 | m | 66-70 | left | rectosigmoid |  | 3.2 | MSI |  | y | y | n | n |
| MSH6_08 | f | 66-70 | left | rectosigmoid | moderate | 2.3 | MSI | MSH6- | y | n | y | n |
| MSH6_09 | m | 56-60 | right |  | poor | 9 | MSI |  | y | y | y | n |
| MSH6_10 | m | 61-65 | right | cecum/ascending colon |  | 6 | MSI | MSH6- | n | y | n | n |
| MSH6_11 | f | 46-50 | right | transverse colon | poor/moderate | 10 | MSI | MLH1+MSH2+MSH6-PMS2+ | n | n | y | n |
| MSH6_12 | f | 46-50 | left | rectosigmoid | moderate | 8 | MSI | MLH1+MSH2+MSH6-PMS2+ | n | n | n | n |
| MSH6_13 | m | 61-65 | left | rectosigmoid | poor |  | MSI | MLH1+MSH2+MSH6- | y | y | y | n |
| MSH6_14 | m |  | left | rectosigmoid | moderate |  | MSI | MLH1+MSH2+MSH6-PMS2+ | y | y | y | n |
| MSH6_15 | f | 71-75 | left | rectosigmoid | moderate |  | MSI | MLH1+MSH2+MSH6-PMS2+ | n | n | y | n |
| MSH6_16 | f | 81-85 | left | rectosigmoid | moderate | 2 | MSI | MSH6- |  | y | n | n |
| MSH6_17 | f | 61-65 | right | cecum/ascending colon |  | 4.5 | MSI | MLH1+MSH2+MSH6-PMS2+ | y | n | y | n |
| MSH6_18 | m | 51-55 | left | rectosigmoid | moderate | 2 | MSI | MLH1+MSH2+MSH6-PMS2+ | n | n | n | n |
| MSH6_19 | f | 46-50 | left | rectosigmoid | moderate |  | MSI | MSH6- | y | n | y | n |
| MSH6_20 | m | 61-65 | right | transverse colon | moderate |  | MSI | MLH1+MSH2-MSH6-PMS2+ | y | y | y | n |
| MSH6_21 | f | 41-45 | left | rectosigmoid | moderate/well | 5.5 | MSI | MLH1+MSH2-MSH6-PMS2+ | n | y | n | n |
| MSH6_22 | m | 41-45 | right | transverse colon | poor | 7 | MSI | MLH1+MSH2+MSH6- | y | y | y | n |
| MSH6_23 | m | 66-70 | right | cecum/ascending colon | moderate/well | 3.5 | MSI |  | y | y |  |  |
| MSH6_24 | f | 61-65 | right | cecum/ascending colon | moderate | 7 | MSI | MLH1+MSH2+MSH6- | y | y | y | n |
| MSH6_25 | m | 46-50 | right | cecum/ascending colon | poor/moderate | 6 | MSI |  | y | y |  | n |
| MSH6_26 | f | 71-75 | right | cecum/ascending colon | moderate | 6 | MSI | MSH6- | y | y | y | n |
| MSH6_27 | m | 31-35 | left | rectosigmoid |  | 3 | MSI | MSH6+ | y | n | y | y |
| MSH6_28 | m | 66-70 |  |  | moderate |  | MSI | MSH6- | y | y | y | n |
| MSH6_29 | m | 75-80 | right | appendix | well | 5 | MSI | MLH1+MSH2+MSH6-PMS2+ | y | n | y | n |
| MSH6_30 | f | 41-45 | right | cecum/ascending colon | moderate | 4 | MSI | MSH6- | y | n | y | n |
| MSH6_31 | f | 46-50 | left | descending colon | poor |  | MSI | MSH6- | n | n | n | n |
| MSH6_32 | m | 71-75 | left | rectosigmoid | moderate | 4.5 | MSI |  | y | y | y | n |
| MSH6_33 | m | 46-50 | left | rectosigmoid | poor/moderate | 7 | MSI |  | n | y | y | n |
| MSH6_34 | m | 75-80 | right | cecum/ascending colon | moderate | 6 | MSI | MLH1+MSH2+MSH6-PMS2+ | y | y | y | n |
| MSH6_35 | m | 36-40 | left | rectosigmoid | moderate | 5.5 | MSI | MLH1+MSH6-PMS2+ | y | y | n | n |
| MSH6_36 | f | 75-80 | left | rectosigmoid | moderate | 2.5 | MSI | MSH6- | n | y | y | n |
| MSH6_37 | m | 66-70 | right | cecum/ascending colon | moderate | 8 | MSI | MSH6- | n | n | y | n |
| MSH6_38 | m | 66-70 | right |  | moderate/well | 2.7 | MSS | MLH1+MSH2+MSH6-PMS2+ | y | y | y | n |
| MSH6_39 | m | 71-75 |  |  | moderate/well | 1.3 | MSS | MSH6+ | n | n | n | n |
| MSH6_40 | m | 81-85 | left | descending colon | moderate/well | 4.5 | MSI | MSH6- | y | y | y | n |
| PMS2_01 | m | 41-45 | left | rectosigmoid | moderate/well | 5.7 | MSI | MLH1+MSH2+MSH6+PMS2- | y | y | n | n |
| PMS2_02 | m | 66-70 | left | descending colon |  | 7 | MSI | MLH1+MSH2+MSH6+PMS2- | n | n | y | n |
| PMS2_03 | m | 66-70 | right |  | moderate/well |  | MSI |  | n | y | n | n |
| PMS2_04 | f | 61-65 | right | cecum/ascending colon | moderate/well | 2.5 | MSI | MLH1+MSH2+MSH6+PMS2- | y | y | y | y, but lower than stroma |
| PMS2_05 | f | 51-55 | left | rectosigmoid |  | 4.1 | MSI |  | y | y | y | n |
| PMS2_06 | m | 56-60 | left | rectosigmoid | well | 5 | MSS | MLH1+MSH2+MSH6+ | y | y | y | n |
| PMS2_07 | f | 26-30 | left | rectosigmoid | poor | 5.4 | MSI | MLH1+MSH2+MSH6-/+PMS2+ | y | y | y | n |
| PMS2_08 | f | 36-40 | right | cecum/ascending colon | moderate/well | 1.5 | MSI |  | n | n | y | n |
| PMS2_09 | f | 46-50 | right | cecum/ascending colon | moderate |  | MSI |  | n | n | y | n |
| PMS2_10 | m | 61-65 | left | rectosigmoid | poor/moderate | 3.5 | MSI |  | y | y | n | n |
| PMS2_11 | m | 41-45 | right | cecum/ascending colon | moderate | 6 | MSI |  | y | y | y | n |
| PMS2_12 | m | 36-40 | right | cecum/ascending colon | moderate | 5 | MSI | MLH1+MSH2+MSH6+ | y | y | n | n |
| PMS2_13 | m | 51-55 | left | rectosigmoid | moderate | 5 | MSI |  | y | y |  | n |
| PMS2_14 | f | 46-50 | left | rectosigmoid | moderate | 1.5 | MSS |  | y | y | y | n |
| PMS2_15 | m | 36-40 | right | cecum/ascending colon | poor | 5 | MSI | PMS2- | n | n | n | n |
| PMS2_16 | f | 51-55 | right | cecum/ascending colon | moderate | 3.5 | MSI | MLH1+MSH2+MSH6-PMS2- | n | n | y | n |
| PMS2_17 | m | 46-50 | right | transverse colon | moderate/well |  | MSI |  | y | y |  | n |
| PMS2_18 | f | 56-60 | right | cecum/ascending colon | moderate/well | 3.5 | MSI |  | n | n | y | n |
| PMS2_19 | f | 56-60 | left | rectosigmoid | poor/moderate |  | MSS |  | y | n | n | n |
| PMS2_20 | f | 41-45 | right |  | moderate | 5 | MSI | PMS2- | n | n | n | n |
| PMS2_21 | f | 41-45 | right | cecum/ascending colon | moderate | 9 | MSI |  | n | n | y | n |
| PMS2_22 | f | 41-45 | right |  |  |  | MSI |  | y | y | y | n |
| *CRC, colorectal cancer; f, female; IHC, immunohistochemistry; m, male; MMR, mismatch repair; MSI, microsatellite instability; MSS, microsatellite stable; n, no; y, yes* | | | | | | | | | | | | |

| **Supplementary Table 5. Median number of cells per MMR group for each phenotype in the IMC dataset** | | | | | | | | | | |
| --- | --- | --- | --- | --- | --- | --- | --- | --- | --- | --- |
| **Phenotype** | ***MLH1*** | | ***MLH1*-PM** | | ***MSH2*** | | ***MSH6*** | | ***PMS2*** | |
|  | **Median** | **IQR** | **Median** | **IQR** | **Median** | **IQR** | **Median** | **IQR** | **Median** | **IQR** |
| Apoptotic cancer cells | 44.25 | 26.875-97.125 | 37.25 | 28.875-62.5 | 27.5 | 8-45.5 | 44 | 6.75-47.875 | 75 | 19-150.75 |
| B cells | 13.25 | 1.375-60 | 7 | 3.25-15.875 | 6.25 | 0.75-15.625 | 2.25 | 1-14.375 | 2 | 0.25-14 |
| CD103^+^ CD4^+^ T cells | 7.75 | 5.25-12.5 | 14.5 | 10.25-23.25 | 6.5 | 2.375-10.125 | 4.5 | 0.75-9.125 | 28.5 | 5-48 |
| CD103^+^ CD8^+^ T cells | 71 | 27.5-100.375 | 103.75 | 69.625-121.75 | 44.5 | 10.625-81.875 | 18.75 | 8.25-48.75 | 106 | 19.75-117.25 |
| CD103^+^ T cells | 35.75 | 17.125-60.25 | 18.25 | 10-48.125 | 4.5 | 2.25-11.25 | 4.75 | 1.75-8.5 | 15 | 7-28.5 |
| CD11b^+^ cells | 175.75 | 84-230.875 | 93.25 | 35.625-120.625 | 40 | 16.25-140.5 | 160.25 | 25.5-254.5 | 73 | 59.75-100.25 |
| CD204^+^ CD163^+^ macrophages | 185 | 139.625-264 | 164.5 | 137.625-203.25 | 188 | 168.25-205.125 | 156 | 124.75-196.25 | 155 | 125.25-224 |
| CD204^+^ macrophages | 43.5 | 28-74.625 | 39.5 | 30.375-52.625 | 92 | 28.5-122.5 | 24.5 | 10-66.75 | 35 | 11.25-43 |
| CD4^+^ T cells | 214.25 | 143.875-305.375 | 237 | 186.625-296.25 | 194.25 | 159.25-268.5 | 117 | 66.875-278.625 | 263 | 61.5-302.25 |
| CD8^+^ T cells | 127 | 90-183.25 | 187.75 | 118.125-222.25 | 90 | 31.875-143.75 | 57.5 | 23.375-77.875 | 157 | 25.5-170.5 |
| D2-40^+^ fibroblasts | 309.75 | 258.375-400 | 201.25 | 190.375-223.5 | 295 | 268.25-334.75 | 385.75 | 319.125-529.75 | 292 | 146.5-317.25 |
| Dendritic cells | 41.25 | 18.875-91.875 | 23.75 | 14-27.25 | 21.5 | 12.5-39 | 9 | 7.625-23.25 | 12 | 4-30.75 |
| Fibroblasts | 1516 | 1223.625-1642.75 | 830 | 582.75-998.75 | 911.25 | 726.25-1287 | 888.25 | 412.25-1317 | 566 | 398.5-694.75 |
| GD T cells | 4.25 | 0.375-12.625 | 4.75 | 2.375-9.125 | 1 | 0.5-4.5 | 0 | 0-2.375 | 7 | 1.75-14.5 |
| Granulocytes | 157.75 | 132.75-172.5 | 113.25 | 12.5-126.875 | 86 | 54.5-126.625 | 55 | 26.625-78.25 | 143 | 79.25-202.5 |
| IDO^+^ cancer cells | 8.25 | 2.875-108.625 | 5.75 | 0.875-55.5 | 0.5 | 0-3.75 | 3 | 0-19.375 | 3 | 0-12.5 |
| Innate lymphoid cells | 39.75 | 12.375-77.875 | 30 | 16.375-42.625 | 20 | 11.875-33.25 | 16.5 | 14.375-29.125 | 19 | 11.5-38.75 |
| Lymph vessel | 22.25 | 18.5-31.625 | 19.75 | 15.375-33 | 19 | 6.375-44.25 | 38.75 | 13.5-44.375 | 37 | 9.5-42 |
| Monocytes | 49.5 | 43.125-79.75 | 50.75 | 42.25-62.375 | 43.5 | 36-82.75 | 46 | 36.75-73.125 | 42 | 31.5-47.5 |
| Other macrophages | 49.25 | 29.25-72.75 | 29.5 | 15.875-39.125 | 34.5 | 10.5-58.25 | 26.75 | 13.75-40.5 | 30.5 | 25.5-43.5 |
| Plasma cells/plasmablasts | 133 | 56.375-420.125 | 146.75 | 115.75-235 | 224.5 | 56-269 | 151 | 62.625-265.25 | 61 | 44-178.5 |
| Proliferating cancer cells | 626.5 | 380.625-759.875 | 708.75 | 608.625-931.375 | 295.75 | 149.75-441.875 | 585 | 202-832.5 | 659.5 | 425-842 |
| p16^+^ cancer cells | 80.5 | 47.75-197.25 | 28.75 | 8.375-80.75 | 56 | 16.5-121.125 | 119.5 | 11-229.25 | 100 | 10.75-231.5 |
| T cells | 562.25 | 347-741.125 | 574.25 | 531.625-668.125 | 389.5 | 230.375-529.25 | 269 | 204-559.75 | 650.5 | 153.75-701 |
| Tregs | 56.75 | 19.875-81 | 46.25 | 38.625-68.125 | 30.5 | 18-45.375 | 19 | 14.875-37.125 | 35 | 12.25-81.5 |
| Cancer cells | 1386.5 | 924.5-1556.375 | 972.5 | 782.875-1644.75 | 1015.5 | 314.75-1456 | 1167.25 | 801.375-2501.375 | 1200 | 841.25-1804.75 |
| Vessel | 219.75 | 172-271.875 | 206.5 | 138.375-234.625 | 203 | 173.5-264.25 | 196.5 | 143.75-276.375 | 140 | 92.5-175.5 |
| *IMC, imaging mass cytometry; IQR, interquartile range; MLH1-PM, MLH1 promotor hypermethylation; MMR, mismatch repair* | | | | | | | | | | |

| **Supplementary Table 6. Median number of cells per MMR group for each phenotype in the mIF and IHC datasets** | | | | | | | | | | | |
| --- | --- | --- | --- | --- | --- | --- | --- | --- | --- | --- | --- |
| **Phenotype** | | ***MLH1*** | | ***MLH1*-PM** | | ***MSH2*** | | ***MSH6*** | | ***PMS2*** | |
|  |  | **Median** | **IQR** | **Median** | **IQR** | **Median** | **IQR** | **Median** | **IQR** | **Median** | **IQR** |
| **All CD4^+^ T cells** | | 195,8445 | 161,813125-248,0635 | 175,1257 | 153,0006-201,46955 | 209,0633 | 186,78565-248,4854 | 207,8575 | 178,05415-252,2635 | 214,8759 | 191,2508-245,204125 |
|  | CD4^+^ T cells | 137,82375 | 95,76601-184,688275 | 123,7505 | 94,781625-151,8756 | 157,3705 | 128,391125-207,7508 | 130,59425 | 104,710625-167,602225 | 137,7193 | 101,8129-181,266325 |
|  | CD4^+^ CD103^+^ T cells | 10,845218 | 7,0744035-16,734445 | 11,43755 | 6,8437775-18,281325 | 16,593815 | 10,581189-40,7255275 | 22,50009 | 11,9062975-41,765795 | 15,65631 | 7,50003025-34,312635 |
|  | CD4^+^ CD103^+^ GZMB^+^ PD1^+^ T cells | 7,203751 | 3,750015-15,70319 | 6,750027 | 2,5312605-12,8438 | 3,937516 | 2,1492865-8,10528225 | 5,906274 | 3,640389-9,6562885 | 7,3125295 | 4,5468935-13,8281825 |
|  | CD4^+^ GZMB^+^ T cells | 17,34382 | 12,421925-27,515735 | 14,06256 | 9,468788-23,15634 | 16,04995 | 9,37503725-26,5388525 | 13,781305 | 8,135902-19,546955 | 16,031315 | 11,671925-25,39618 |
|  | CD4^+^ PD1^+^ T cells | 9,424743 | 3,42188875-18,336695 | 8,062532 | 3,937516-18,37507 | 13,4063 | 6,70315175-21,75009 | 14,53131 | 8,3430325-24,578225 | 16,48357 | 9,1406615-27,1876075 |
| **All CD8^+^ T cells** | | 164,7652 | 94,5941275-258,776775 | 104,2504 | 27,28136-186,75075 | 166,50065 | 110,297275-252,759275 | 135,56305 | 66,7971425-214,6884 | 169,2967 | 140,46195-235,922775 |
|  | CD8^+^ T cells | 83,625335 | 57,7033575-183,830575 | 56,62523 | 11,343795-117,563 | 114,1914 | 72,65654-195,54075 | 74,344045 | 29,859495-137,719275 | 77,89064 | 55,82835-147,328725 |
|  | CD8^+^ CD103^+^ T cells | 16,312565 | 9,0469115-29,378365 | 19,31258 | 6,468776-27,00011 | 20,53133 | 7,3594045-29,6562475 | 17,8986 | 6,71036325-26,484485 | 28,09736 | 16,2656875-36,513965 |
|  | CD8^+^ CD103^+^ GZMB^+^ PD1^+^ T cells | 6,468776 | 4,78126925-8,292342 | 12,75005 | 5,06252-19,68758 | 8,812535 | 3,750015-15,7886875 | 9,906289 | 4,83715875-26,390735 | 21,014505 | 12,03927-39,1407825 |
|  | CD8^+^ GZMB^+^ T cells | 14,06256 | 3,46876425-28,7344925 | 4,312517 | 1,312505-10,31254 | 13,218805 | 6,93752825-26,305605 | 9,20846 | 3,70313975-22,07821 | 13,593805 | 7,8633125-35,0157625 |
| **All CD204^+^ macrophages** | | 67,21317 | 52,68771-79,54719 | 77,56281 | 51,89083-95,81138 | 62,22681 | 53,3232725-73,80119 | 64,223025 | 57,09398-85,0550275 | 51,28146 | 40,31266-69,65653 |
|  | CD204^+^ HLA-DR^+^ macrophages | 23,6906 | 17,34382-28,75255 | 23,95322 | 15,84381-41,06266 | 25,91481 | 13,7930275-36,2305375 | 23,27353 | 15,681335-39,00016 | 19,31258 | 15,09381-22,57122 |
|  | CD204^+^ HLA-DR- macrophages | 38,5314 | 34,24401-55,76347 | 50,0627 | 31,12512-63,04713 | 37,933415 | 28,6758925-45,6868375 | 40,85543 | 30,5391875-51,3891225 | 36,36473 | 31,3595-45,03899 |
| **All granulocytes** | | 73,03154 | 56,32485-88,38573 | 74,57842 | 57,37523-110,1567 | 60,072055 | 48,34462-74,57467 | 71,87841 | 49,756565-85,8167475 | 91,44547 | 55,07835-115,3603 |
|  | CD15^+^ HLA-DR^+^ granulocytes | 7,828156 | 3,41592-18,45877 | 12,70318 | 9,375038-19,0782 | 20,534475 | 15,4858525-31,0079375 | 13,804745 | 7,558624-25,265725 | 10,15232 | 6,984403-21,98446 |
|  | CD15^+^ HLA-DR- granulocytes | 67,68777 | 47,67207-75,01269 | 59,71899 | 44,43768-94,59413 | 39,718155 | 26,9499375-49,034915 | 52,673205 | 41,04766-69,5675075 | 60,43429 | 42,8908-93,04725 |
| **γδ T cells** | | 7.63 | 2.05-20.77 | 4.11 | 0.37-7.95 | 7.49 | 2.83-11.30 | 4.83 | 2.69-15.27 | 8.12 | 4.05-29.71 |
| *IHC, immunohistochemistry; IQR, interquartile range; mIF, multiplex immunofluorescence MLH1-PM, MLH1 promotor hypermethylation; MMR, mismatch repair* | | | | | | | | | | | |
